# How can we enhance HIV Pre Exposure Prophylaxis (PrEP) awareness and access?: Recommendation development from process evaluation of a national PrEP programme using implementation science tools

**DOI:** 10.1101/2022.06.09.22276189

**Authors:** Paul Flowers, Jennifer MacDonald, Lisa McDaid, Rak Nandwani, Jamie Frankis, Ingrid Young, John Saunders, Dan Clutterbuck, Jenny Dalrymple, Nicola Steedman, Claudia Estcourt

**Affiliations:** School of Psychological Sciences & Health, University of Strathclyde, Glasgow, UK, 16 Richmond Street, Glasgow G1 1XQ; School of Health & Life Sciences, Glasgow Caledonian University, Glasgow, UK, Cowcaddens Road, Glasgow, G4 0BA; Institute for Social Science Research, The University of Queensland, Brisbane, Australia, Brisbane St Lucia, QLD 4072; College of Medical, Veterinary & Life Sciences, University of Glasgow, Glasgow, UK, University Avenue, Glasgow, G12 8QQ; School of Health & Life Sciences, Glasgow Caledonian University, Glasgow UK, Cowcaddens Road, Glasgow, G4 0BA; Centre for Biomedicine, Self & Society, University of Edinburgh, Edinburgh, UK, Usher Institute, Teviot Place, Edinburgh, EH8 9AG; Institute for Global Health, University College London, London UK, Mortimer Market Centre, London, WC1E 6JB; HPA Health Protection Services, Public Health England, London, UK, 61 Colindale Avenue, London, NW9 5EQ; Chalmers Sexual Health Centre, NHS Lothian, Edinburgh, UK, 2A Chalmers Street, Edinburgh, EH3 9ES; Chief Medical Officer Directorate, Scottish Government, Edinburgh, UK, St Andrew’s House, Regent Road, Edinburgh, EH1 3DG

**Keywords:** HIV, PrEP, Implementation Science, Awareness, Access, Behaviour Change Wheel, Theoretical Domains Framework, Deductive Thematic Analysis, Socio-ecological Model, Qualitative

## Abstract

**Objectives:** HIV Pre-Exposure Prophylaxis (PrEP) is a highly effective biomedical intervention for HIV prevention and is key to HIV transmission elimination. However, implementation is challenging. We identified barriers and facilitators to PrEP awareness and access during the roll out of Scotland’s national PrEP programme to develop recommendations for future provision.

**Design:** Multi-perspectival qualitative approach incorporating implementation science tools.

**Setting:** Sexual health services and sexual health/HIV community-based organisations (CBOs) in Scotland.

**Participants:** Semi-structured telephone interviews and focus groups with geographically and demographically diverse patients seeking/using/declining/stopping PrEP (n=39), sexual healthcare professionals (n= 54), CBO users (n=9) and staff (n=15).

**Analysis:** Using deductive thematic analysis we mapped barriers and facilitators to PrEP awareness and access. We then applied the Theoretical Domains Framework, Behaviour Change Wheel, and Behaviour Change Technique Taxonomy to analyse barriers and facilitators to generate targeted solutions. Finally, we applied APEASE criteria, expert opinion, and the socio-ecological model to synthesise and present multi-levelled and interdependent recommendations to enhance implementation.

**Results:** Barriers and facilitators were multifaceted, relating to the macrosocial (e.g., government, service ecology), the mesosocial (e.g., values and practices of organisations and dynamics and norms of communities) and the microsocial (peer influence). We derived 28 overarching recommendations including: incentivising organisations to share expertise, addressing future generations of PrEP users, expanding the reach of PrEP services, cascading effective service innovations, changing organisational cultures, instigating and managing novel outreach, establishing monitoring systems, supporting diverse PrEP users, providing training addressing awareness and access to professionals, and development of “PrEP champions” within a range of organisations.

**Conclusion:** Improving awareness and access to PrEP sustainably will require intervention across the whole system, changing policy and practice, organisations and their cultures, communities and their social practices, and individuals themselves. These evidence-based recommendations will prove useful in extending the reach of PrEP to all who could benefit.

**Article Summary:** *Strengths and limitations of this study:*

- We used novel methods and a rigorous study design to create auditable evidence-based and theoretically informed recommendations, moving beyond simple thematic analysis or sole use of expert opinion
- The recommendations are built upon multi-perspectival qualitative data from diverse stakeholders and varied expert opinions.
- Where meta-analyses or meta-syntheses of implementation studies are not available, we offer a structured, practical, evidence-based approach to generating recommendations.
- Limitations include the sole reliance on qualitative insights and our focus on a single national context (Scotland) in the early years of programme delivery.

## INTRODUCTION

Oral HIV pre-exposure prophylaxis (PrEP), involves taking antiretroviral medication to prevent HIV acquisition. PrEP has an established and increasingly important role in HIV prevention [1, 2] and will be a key component of HIV transmission elimination strategies. However, major challenges to its implementation remain.[3-5] There are inequities in PrEP roll out globally,[6] and uptake varies across minoritised populations such as men who have sex with men, women of colour, people who inject drugs, migrants and trans people.[6-9] International models of PrEP provision are diverse and include provision by HIV specialists, sexual health specialists, family medical practices, and outreach programs.[6]

Current evidence on PrEP implementation draws on a broad range of disciplinary lenses and takes disparate focal points.[10] Specific approaches to conceptualising PrEP implementation include Liu’s PrEP cascade [11] the PrEP continuum of care [12] and the PrEP care continuum.[13] These conceptualisations, and associated literature, are intended to assist with framing, auditing and responding to PrEP implementation challenges. However, the mechanisms by which these high-level conceptualisations enhance future PrEP implementation remain opaque and they fail to provide detailed recommendations for action. Emerging literature, primarily from the USA, also highlights the multi-levelled nature of barriers and facilitators to PrEP implementation, such as individual, relational, community and policy-related influences.[14, 15] To date, no studies have taken a systematic approach to addressing these multi-levelled influences on implementation. We address this gap directly, drawing upon experience of the first two years of Scotland’s national PrEP programme, to develop evidence-based and theoretically informed recommendations to enhance PrEP implementation. In parallel papers, using the same datasets (published elsewhere) we explore issues of PrEP uptake and initiation (Estcourt et al., in preparation) and adherence and retention (MacDonald et al., in preparation). Here we focus on PrEP awareness and access, the first critical steps towards extending the advantages of PrEP to all who may benefit.[16]

### Research questions

1. What were the key barriers and facilitators to PrEP awareness and access within Scotland’s PrEP programme?
2. What evidence-based and theoretically informed recommendations to enhance PrEP awareness and access can be made?

## METHODS

### Overview

Increasing awareness and access to PrEP requires consideration of settings, contexts, participants, actions and targets.[17] The breadth of these considerations requires a comprehensive methodological approach making use of a range of analytic techniques and explanatory frameworks.[18] We conducted a multi-perspectival qualitative study, using semi-structured telephone interviews and in-person focus groups with geographically and demographically diverse participants.

We used thematic analysis as a starting point then applied implementation science tools to systematically develop recommendations for enhancing implementation.

The four tools were; the theoretical domains framework (TDF),[19] the behaviour change wheel (BCW),[20] the behaviour change technique taxonomy (BCTT),[21] and the APEASE (Acceptability, Practicability, Effectiveness, Affordability, Side-effects, and Equity) criteria.[20] We also use Bronfrenbrenner’s Socio-ecological model [22] as an explanatory model (see Figure 1).

**Figure 1.**
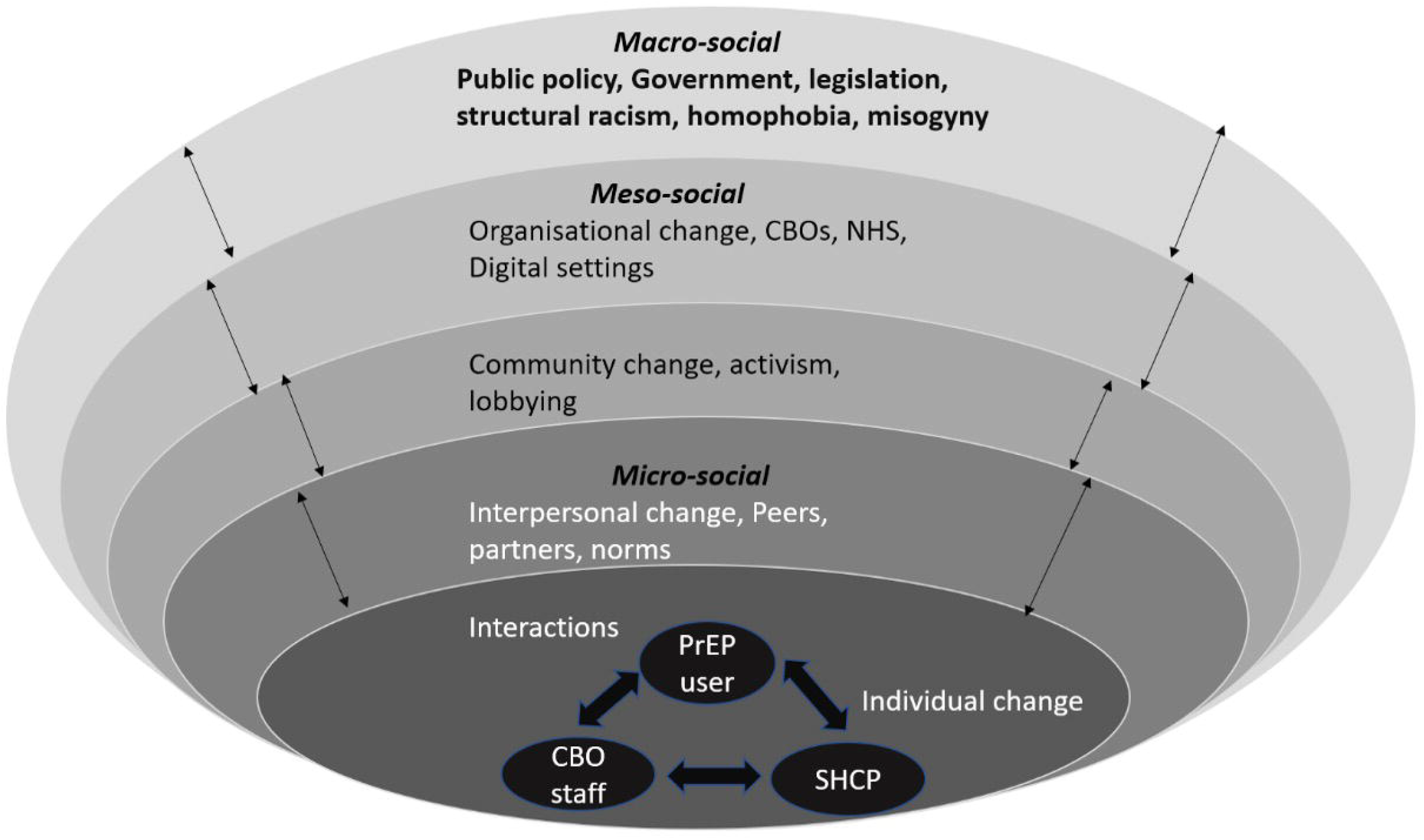
Inter-dependent, multi-levelled, dynamic upstream and downstream determinants of PrEP awareness and access (using the socioecological model*) CBO=community based organisation, HCP=sexual health care professional, NHS=national health service *Adapted from Bronfenbrenner, U. (1979). The ecology of human development: Experiments by nature and design. Harvard University Press [22]. Figure 1 shows the interdependent, multi-levelled determinants of PrEP awareness and access. This recognises that many drivers of health operate far beyond the individual. Critically, causal influence ripples both down-stream and upstream. Equally the same causal influences are felt differentially and simultaneously across multiple levels.

### Context

Scotland was one of first countries worldwide to implement a national PrEP programme.[23] The programme was implemented in specialist sexual health services and was made available to Scottish residents who met HIV risk-based eligibility criteria.[23] All clinical care and medication were provided free of charge in concordance with all NHS delivered sexual health care in Scotland. The program was successful in reducing HIV incidence,[24] but has primarily served gay, bisexual and other men who have sex with men (GBMSM) with less impact on other groups and has only identified a minority of individuals with ‘potentially PrEP preventable’ HIV infection.[25] There was no large-scale awareness intervention when the programme was launched to avoid overwhelming clinic-based services due to the anticipated high early demand for PrEP. Instead, awareness-raising occurred within clinics and via community-based organisations (CBO).[26]

### Data collection

#### Recruitment

We purposively sampled diverse key stakeholders across Scotland. HCPs offered sexual health clinic attenders the opportunity to take part in the study across the four urban Health Boards (large organisations within Scotland responsible for their population’s health) in which over 90% of PrEP in Scotland is prescribed. CBO service users were invited to participate in the study via interactions with CBO staff. We recruited these CBO staff, others working within the same CBOs, and HCPs from all 14 Health Boards by email invitation to specific staff.

#### Data collection

All participants provided informed verbal or written consent immediately prior to the interviews and group discussions. We collected data with the aid of a guide that included open-ended questions designed to explore participants’ experiences and perceptions of the implementation of PrEP. Where possible within the group discussions, dialogue between participants was encouraged rather than between facilitators and participants. All participants talked from their own and others’ perspectives; data were taken at face value. Clinic attenders were offered a £30 shopping voucher as reimbursement for their time. Data collection was led by JM, with input from experienced qualitative researchers, PF, IY, and JF. JM, PF, IY, and JF reviewed and discussed early transcripts for quality assurance purposes. Interviews and group discussions were audio-recorded, transcribed verbatim, anonymised, and imported into NVivo.

#### Participants

In total, 117 participants took part in in-depth, semi-structured telephone interviews (n=71) or group discussions (n=46) between September 2018 and July 2019. The sample comprised: 39 clinic attenders; 54 healthcare professionals (HCPs); 9 CBO service users; and 15 CBO staff from across Scotland. Group discussions included one type of stakeholder only. All CBOs had an HIV prevention remit and served GBMSM, transgender, and Black African communities.

Clinic attenders were either using (n=23, 59%), stopping (n=6, 15%), declining (n=5, 13%), or assessed as ineligible for (n=5, 13%), PrEP. Ages ranged from 20 to 72 years with just over half (n=21, 54%) aged between 25 and 34 years. All self-identified as men, the majority of whom (n=34, 87%) were cisgender, and gay and bisexual men who have sex with men (GBMSM). Almost all clinic attenders reported their ethnicity as ‘White British’ (n=31, 80%) or ‘Other White’ (n=7, 18%). Most reported a university degree as their highest level of education (n=26, 67%) and that they were currently working (n=34, 87%). The area of residence of clinic attenders reflected a mix of affluence and deprivation.

HCPs were all involved in PrEP implementation and included specialist sexual health clinicians (doctors and nurses of varying seniority and experience), health promotion officers, a midwife, and a clinical secretary with responsibility for PrEP administration. They worked in a mix of rural (n=12, 22%), semi-rural / urban (n=8, 15%), or urban (n=34, 63%) settings. Demographic details of the CBO staff were not recorded and CBO service users were all of Black African ethnicity, predominantly women, and not using PrEP.

### Patient and Public Involvement Statement

Public and patient involvement was central to the project and its iterative development, yet this was not limited to a formal PPI workstream. The project developed from and was continuously shaped by collaborations between those involved with making PrEP policy, community-based organisations working directly with PrEP users, clinicians and researchers in the PrEP field. PrEP users were also part of the research team.

### Data analysis

#### Preliminary conceptual work

Given the complexity and span of the potential ways in which PrEP awareness and access could be improved, it was important to focus our main analyses where they were needed most (i.e., where future interventions could enhance PrEP awareness and access). To achieve this, we developed a series of visualizations to conceptualise PrEP awareness and access in terms of the sequential actors, actions, settings and processes involved. We drew on UK policy documents relating to PrEP (see Figure 2 in supplementary files). A group of five experts with experience of developing and implementing interventions to increase awareness and access within their professional/organisational roles used the visualizations to help select priority areas which they felt were most important. These predominantly focussed on actions to support the eventual PrEP user, and linked to CBOs and journeys to, and through, NHS clinical settings.

Priority areas were: (1) Potential PrEP users acquiring knowledge of HIV and its transmission risks in addition to acquiring knowledge of PrEP itself; (2) Potential PrEP users acquiring accurate perceptions of their PrEP candidacy; (3) PrEP users discussing PrEP with others; (4) CBO staff raising issues of PrEP with key communities; (5) HCPs acquiring knowledge of PrEP; (6) HCPs engaging potential PrEP users with PrEP; (7) Sexual health services providing access to PrEP; and (8) Potential PrEP users accessing sexual health services and PrEP care therein.

These eight priority areas were then used to frame the analysis of barriers and facilitators to PrEP awareness and access.

#### Barriers and facilitators to PrEP awareness and access (Research Question 1)

We conducted deductive thematic analysis.[27] Taking each of the eight priority areas separately and treating each as the focus of an independent behavioural analysis we used relative frequency of barriers and facilitators and their amenability to change as our measure of salience. Details of all salient barriers and facilitators are available within supplementary file B. We then synthesised the salient barriers and facilitators within the socioecological model (Figure 1). This work was led by PF and JM and validated with the wider team. This synthesis step was done to manage the volume and multi-levelled complexity of the salient barriers and facilitators for ease of communication with diverse audiences, and to emphasise the inter-dependencies between the individual salient barriers and facilitators.

#### Creation of the recommendations (Research Question 2)

We initially used the TDF to theorise each salient barrier and facilitator (mapping their role as causal influences on implementation across 14 theoretical domains). The TDF is a meta-theoretical tool, synthesising multiple attempts to theorise causal influences on implementation behaviours.[19] Next, we used the BCW [20] to build on the TDF analysis to systematically generate novel intervention elements. The BCW is another tool that brings a meta-perspective to build new intervention content that is both theorised and matched to demonstrable need. As such we used the BCW to suggest corresponding intervention content for the salient barriers and facilitators. These intervention functions were described in further detail using the BCTT (a tool which allows behaviour change intervention content to be explicated in the most granular way possible). JM led these analyses and generated initial recommendations. These were double-checked for accuracy, validity, and credibility by PF. Both JM and PF were trained in using the BCTT. Disagreements were reviewed until consensus was reached.

The initial recommendations were further scrutinized and sense-checked by team members with direct responsibility for delivering and planning PrEP services using the APEASE criteria (Acceptability, Practicability, Effectiveness, Affordability, Side-effects, and Equity) to remove, or adapt any recommendations that were not acceptable, practicable, likely to be effective, likely to be affordable, have side effects or lead to inequity (APEASE criteria, Michie et al., 2014).[20]

To assist with the volume and multi-levelled complexity of the results of the analyses, we synthesised the final recommendations by de-duplicating and putting ‘like with like’ for ease of communication with diverse audiences, and to emphasise the inter-dependencies between the recommendations. This stage was conducted by PF, JM and CSE and checked by the wider team. This approach ensured there was an audit trail available for each of the recommendations. Please see Supplementary file B for the detailed analyses.

## RESULTS

### 1. What were the key barriers and facilitators to PrEP awareness and access?

There was marked heterogeneity in the causal influences shaping PrEP awareness and access with dynamic interplay between a range of upstream and downstream factors. Many findings related to both awareness and access (Table 1).

**Table 1.** An overview of key barriers and facilitators to increasing PrEP awareness and access from the deductive thematic analysis-with illustrative data extracts.

| Typical socioecological model elements * | Key barriers | Key facilitators |
| --- | --- | --- |
|  | **The macrosocial level** |  |
| **Public Policy** | **Enduring structural factors constrain awareness and access to PrEP** (e.g., poverty, racism).<br>**Limited resource and capacity constrained access to PrEP:** *“even the simple fact of releasing people from clinics to do any training was a challenge.” (HCP)*<br>**On-going intractable stigmas limited access to PrEP** (e.g., homophobia, negative sex attitudes)<br>**Historic lack of sexual health education limited awareness and access to PrEP:** *“Because nothing like this was talked about at school. And it's cost lives, the long and the short of it, we've lost people due to ignorance, to lack of education.” (PrEP user)* | **Political will enabled access to PrEP for many:**<br>*“It was a tiny bit chaotic, because the timescale was not of my choosing [.....] and seemed to be be overtly political, because there was a real drive to make Scotland first.” (HCP)*<br>**Sharing learning across diverse networks enabled awareness and access to PrEP** *“There's ongoing dialogue between ourselves and our frontline workers, and the people who commission the service. (CBO staff working with GBMSM)* |
|  | **The mesosocial level** |  |
| **Organisational** | **Struggling to adapt sexual health promotion skills and experience can limit awareness and access to PrEP:** *“I think it's fair to say that the NHS and the third sector are going through a period of kind of cultural change in order to be able to accommodate this new element of messaging. I mean, the landscape has changed enormously.” (CBO staff working with GBMSM)*<br>**Culturally uninformed services reduced access to PrEP for some under-served communities:** *“...it's a minefield for trans people.” (CBO staff working with trans people)*<br>**Uninformed general health services limited access to PrEP:** *“My own GP at the time knew nothing about PrEP, absolutely nothing at all. So I kind of just gave up on that avenue and realised that the clinic was the best place to talk about it.” (PrEP user)*<br>**Failure to use a wide range of settings to promote PrEP limited access** (e.g., primary care, outreach, community settings)<br>**Overstretched sexual health services limited access to PrEP:** *“PrEP is another thing to try and add into an already kind of lengthening* | **Expanding PrEP services beyond sexual health services would increase access to PrEP in the future:** *“for your more straightforward patients, is the ability for other services such as, I guess primary care, to do some of the follow up, and for us to maybe see them every year.” (HCP)*<br>**Culturally informed services increased access to PrEP for some under-served communities** (e.g., those with visibly inclusive services and well trained staff)<br>**Sharing service innovations across the sector increased access to PrEP** (e.g., sharing protocols as they are developed)<br>**The use of nurse prescribers in some sexual health services increased access to PrEP:** *“Cos nurse led services will win hands down. So if a service was planning this and had time, I would say get all your nurses to go through a... some kind of non-medical prescribing.” (HCP)*<br>**Normalising PrEP interactions within sexual health services increased access:** *““When we first started, I was having longer appointments and actually, after a few months, once I'd got to grips with that, we could see maybe a few more people in the PrEP clinic and just because we're* |
|  | *consultation.” (HCP)* | *better now, we’ve got our spiel, we’ve got our way of doing it.” (HCP)* |
| **Community** | **Decisions not to have a mass media intervention limited population-level awareness of PrEP:** “we weren’t out there shouting from the rooftops about PrEP, and we should have been.” (CBO staff working with GBMSM)<br>**Decisions not to have a mass media intervention limited awareness amongst those who could benefit** (e.g., closeted gay men, trans people)<br>**A lack of broad representation in PrEP materials limited awareness and access:** “... it’s not working for the African community, because the messages have to be strong and specific, the communities who can access PrEP. It’s not just about gay men, and that’s the way it’s been. It needs to be a strong voice to say, it’s for everybody.” (CBO staff working with Black African communities) | **The visibility of PrEP in dating Apps facilitated awareness and increased access:** “I use some gay dating apps and a lot of people mentioned on their profile that they use PrEP, and at some point I kind of asked around and I found out that this is something that you could take to prevent you from getting HIV.” (PrEP user)<br>**Changing norms about PrEP within communities increased access** (e.g., new norms about asking about PrEP use)<br>**Peer influence in some communities increased awareness and access to PrEP** (e.g., People talk with friends and networks about PrEP and increase access)<br>**An understanding of the wider benefits of PrEP amongst HCPs increased access** (e.g., HCPs appreciate longer term societal benefits of PrEP) |
| **The microsocial level** |  |  |
| **Interpersonal** | **Written materials about PrEP limited awareness and access:** “The reading uptake among our people. Remember, if you go back to our history, we lived in oral history” (CBO staff working with Black African communities)<br>**Changing communication norms limited awareness and access:** “I mean, the best will in the world, nobody takes written information any more.” (HCP) | Within some communities, social networks and norms enabled awareness and access: “I was aware of people that I’ve known that have had HIV and have had it for quite a long time and been on treatment and been almost survivors or such. So I knew there was a quite a lot of advancement.” (PrEP user) |
| **Individual change** | **HCPs lack of confidence in identifying those who may benefit from PrEP limited access to PrEP:** “Women who are at risk of HIV are probably pretty difficult to identify. I’d say, particularly people who are in a relationship, they’re very difficult to identify” (HCP)<br>**Individual HIV risk perceptions limited access:** “There was a part of me that thought, actually, this is an intervention that only people who are putting themselves at risk need, you know. Someone like me doesn’t need it, because I’m not like that. But that’s silly, of course, that was silly.” (PrEP user) | **Individuals increased PrEP awareness because of community dynamics:** “I use some gay dating apps and a lot of people mentioned on their profile that they use PrEP, and at some point I kind of asked around and I found out that this is something that you could take to prevent you from getting HIV.” (PrEP user) |

\*interdependent and overlapping levels from macro-system to micro-system

Key barriers and facilitators were multi-levelled. At the macrosocial level facilitators included governmental will and effective shared learning across networks. Barriers included structural issues such as poverty and racism, enduring stigmas, the ongoing impacts of inadequate sexual health education. In turn, these macrosocial barriers and facilitators shaped, and were shaped by mesosocial-level issues. For organisations, barriers included difficulties adapting their existing skills and experience to PrEP delivery, the existence of services that lacked cultural competence, not using the full range of available settings to promote PrEP, overstretched NHS and CBO services. Facilitators included expanding professional roles within sexual health services such as involving nurse prescribers, and normalising PrEP interactions within sexual health services. At the community-level, barriers included the limited representation of communities and diversity within available materials focusing on PrEP, and the lack of mass media and social media interventions to promote PrEP. Facilitators included dating apps increasing awareness of PrEP in some communities and changing norms.

At the microsocial level barriers included health care professionals lacking confidence in identifying those who may benefit from PrEP, the constrained modality of communicating about PrEP, (e.g., relying on leaflets), individual HIV risk perception. Facilitators included the role of peer influence; the role of knowledge of wider benefits of PrEP amongst health care professionals; the role of social networks; individual learning from peers within some communities.

### 2. What evidence-based and theoretically informed recommendations to enhance PrEP awareness and access can be made?

Recommendations derived from the barrier and facilitator analysis and application of APEASE criteria are shown below. Table 2 highlights the macrosocial level; Table 3 highlights the mesosocial level; and Table 4 focusses upon the microsocial level.

**Table 2.** Recommendations for improving PrEP awareness and access at the macrosocial level.

| Typical socioecological model elements | Those in decision-making roles/e.g.national leads/commisioners should:- | Those working within sexual health services should:- | Those working within community-based organisations should:- |
| --- | --- | --- | --- |
| **Public Policy change** | **Use political will and momentum** Create and harness political will and momentum to initiate and then maintain appropriately funded PrEP programmes over an appropriate time scale. Key stakeholders could/should? produce a business case detailing the health and economic benefits of PrEP implementation and the likely cost effectiveness of investing in awareness raising to increase access (13, 14, 63, 64).<br>**Maintain networks** Maintain a network of national and local diverse organisations across health and social care and third sector. Focus on ways of sharing learning to enable optimal approaches to increasing awareness and expanding access across time (1,2,3,5, 13,49,70).<br>**Ensure organisations work together** Ensure funding mechanisms for commissioned PrEP work drive innovation and incentivise partnerships between organisations. Commissioning briefs and service level agreements should explicitly incentivise effective partnerships (1,6).<br>**Address new generations of PrEP users early** Ensure age-appropriate and comprehensive sex and relationships education includes issues such as PrEP as HIV prevention (10) within a sex positive ethos which includes the sexual health, social, and cultural needs of all relevant local communities (e.g., Trans communities) | **Drive effective partnerships** Enhance and maintain good connections across the HIV prevention and care sector and other specialist services - both statutory and community: consider joint roles, joint meetings, and creating formal and informal mechanisms to share data (5, 61). | **Drive effective partnerships** Expand and maintain connections with other organisations (both statutory and community). To increase access, actively encourage reciprocal referrals with non-HIV specialist organisations such as those working with addictions, mental health, and refugees (3,5).<br>**Share knowledge and Skills** Champion and foster shared learning across diverse organisations about what has worked to enhance PrEP access and awareness: dedicate focussed meetings to lessons learned from working with diverse vulnerable populations (e.g., GBMSM, Trans, Black African, injecting drug using communities) (8,40, 61, 69).<br>**Support PrEP users** Activate, support and facilitate the representation and involvement of diverse PrEP users and potential PrEP users within all aspects of policy development and subsequent policy changes: focus on recruiting diverse potential and actual PrEP users and provide opportunities for training and on-going support for them (61). |

\* Each recommendations represents an evidence-based and theoretically-informed way to enhance PrEP awareness and access. To stress their interconnectedness they are again presented within the socioecological framework to signal how each level of recommendation shapes the others. In order to aid with key people making practical use of our recommendations they are grouped according to the sector they are directed at (i.e., commissioners, sexual health service providers and CBOs). The numbers in brackets relate to the specific recommendations generated from the detailed analysis of each barrier and facilitator and can be cross-referenced within the supplementary files.

**Table 3.** Recommendations for improving PrEP awareness and access at the mesosocial level.

| Typical socioecological model elements | Those in decision-making roles e.g. national leads/commissioners should:- | Those working within sexual health services should:- | Those working within community-based organisations should:- |
| --- | --- | --- | --- |
| **Organisational change** | **Establish monitoring systems:** Create and maintain monitoring systems to assess the development of networks connecting diverse organisations and the effectiveness of their partnership working: develop and assess outcomes that reflect cross-sectorial work (71).<br>**Incentivise improved organisational cultures:** Develop a badging system that reflects and publicly shares the achievement and ongoing maintenance of inclusive, culturally competent services across the whole network of organisations involved in PrEP awareness and access (85). | **Expand PrEP services off- and online:** Develop and maintain range of service models and providers to make PrEP available in different settings meeting the needs of those from diverse cultural backgrounds (e.g., people who inject drugs) with different levels of health and service literacy. Each service model should incorporate pathways for people with non-complex PrEP needs and for those additional medical complexity. Settings could include sexual health services, primary care, online PrEP clinics, outreach clinics in community settings (74). Ensure nurse-led care and prescribing is expanded (66).<br>**Cascade service innovations:** Develop, maintain, monitor and share a range of innovations concerning awareness and access (e.g., protected spaces for women, nurse led pathways for non-complex PrEP users). Consider paper-based or electronic checklists and flow charts to remind HCPs of PrEP processes, e.g., a protocolled approach to eligibility criteria including nuanced examples (19, 48, 62, 72, 74, 76, 78). Before scale up of service innovations, develop pilots, a range of peer support opportunities and shadowing opportunities for staff to learn how to engage with new systems, patient groups and collectively develop efficient consultations. Ensure a range of formal and informal opportunities are available to consistently cascade changes to PrEP guidance (18, 49, 67), e.g., developing scripts to succinctly and accurately discuss PrEP (68).<br>**Establish monitoring systems:** Develop, maintain and share monitoring systems to measure PrEP uptake, PrEP need, PrEP reach into KVPs, effective organisation learning and the routinisation of innovations (23, 71).<br>**Participate in an enabling an inclusive organisational culture:** Establish, actively maintain and celebrate a positive organisational culture ensuring professionals competencies celebrate holistic and<br>inclusive understandings of sexual health and can achieve increases in wellbeing, reductions in inequalities, racism, heterosexism, transphobia and homophobia. These should be enshrined in the appearance of the services (off- and online, organisational values and mission statements). Ensure physical space, signage, and materials clearly show cultural and gender diversity and are explicit that PrEP is relevant to all individuals who could benefit, not only GBMSM. Organise dedicated time to share learning about what is and is not working well for underserved communities such as Trans men for example (33,43,47,86, 87,88,89). | **Establish monitoring systems :** Develop and maintain monitoring systems to constantly enhance activities that drive effective partnerships across organisations and that support PrEP users and potential users. Ensure trustees, CEOs and all staff prioritise effective partnership work and that this is reflected within key performance indicators (71).<br>**Cascade innovations:** Encourage shared learning across CBOs and communities to build on success (e.g., activism and peer-influence amongst GBMSM) and transfer to other communities (e.g., people who inject drugs, trans-communities, Black African communities) (61). |
| **Community change** | **Establish and sustain multi-levelled awareness raising interventions** Co-ordinate multi-modal, inclusive, awareness raising, PrEP-normalising, PrEP-positive interventions capable of deep reach adopting a ‘needs-based’ terminology. Examples include mass media/social marketing approaches co-produced with opinion leaders, clearly branded materials visibly endorsed by reputable organisations working in partnership. Materials should be hosted and shared through SHS, CBOs, and HIV/PrEP activists’ websites and social media, posters in CBO, SHS, and other health settings, CBO outreach work, message blasts on dating apps and hook-up sites (31,32,35,41,58).<br>Awareness raising interventions should be complemented by peer-led interventions, particularly in relation to underserved communities (40, 44). | **Develop tools that facilitate shared learning** HCPs, CBOs and communities should co-produce resources that address a range of HCP and CBO educational needs about awareness and access. Examples of culturally appropriate language and scripts to help HCPs assess and highlight HIV risk in culturally competent ways with diverse groups would be useful as well as details of how to appropriately motivate PrEP use (20, 27, 28, 34, 42, 44, 45, 68). | **Instigate and manage novel outreach** Working in partnership with SHS, CBOs should explore innovative ways of outreach (drop in, peer support) to sensitively engage key communities (e.g. Black African or Trans communities) (38, 55).<br>**Co-create socially situated PrEP awareness raising materials** CBOs should lead the co-creation of educational approaches that clearly situate PrEP within the socio-economic and cultural factors of PrEP users’ lives. These approaches should normalise sexual health and SHS attendance. They must be disseminated broadly, making use of a wide variety of settings and modalities. Examples could include flyers, posters, short videos, interactional workshops at diverse community venues, drop-in information sessions, peer-led support groups. Materials should be distributed in non-sexual health-specific health services (4,12,22,36,39,52, 69,81).<br>**Co-create socially situated PrEP awareness raising materials** Ensure dedicated materials also address potential PrEP users’ understanding of what to expect within a service offering PrEP and a potential walk through of a typical patient pathway (82). |

\* Each recommendations represents an evidence-based and theoretically-informed way to enhance PrEP awareness and access. To stress their interconnectedness they are again presented within the socioecological framework to signal how each level of recommendation shapes the others. In order to aid with key people making practical use of our recommendations they are grouped according to the sector they are directed at (i.e., commissioners, sexual health service providers and CBOs). The numbers in brackets relate to the specific recommendations generated from the detailed analysis of each barrier and facilitator and can be cross-referenced within the supplementary files.

**Table 4.**
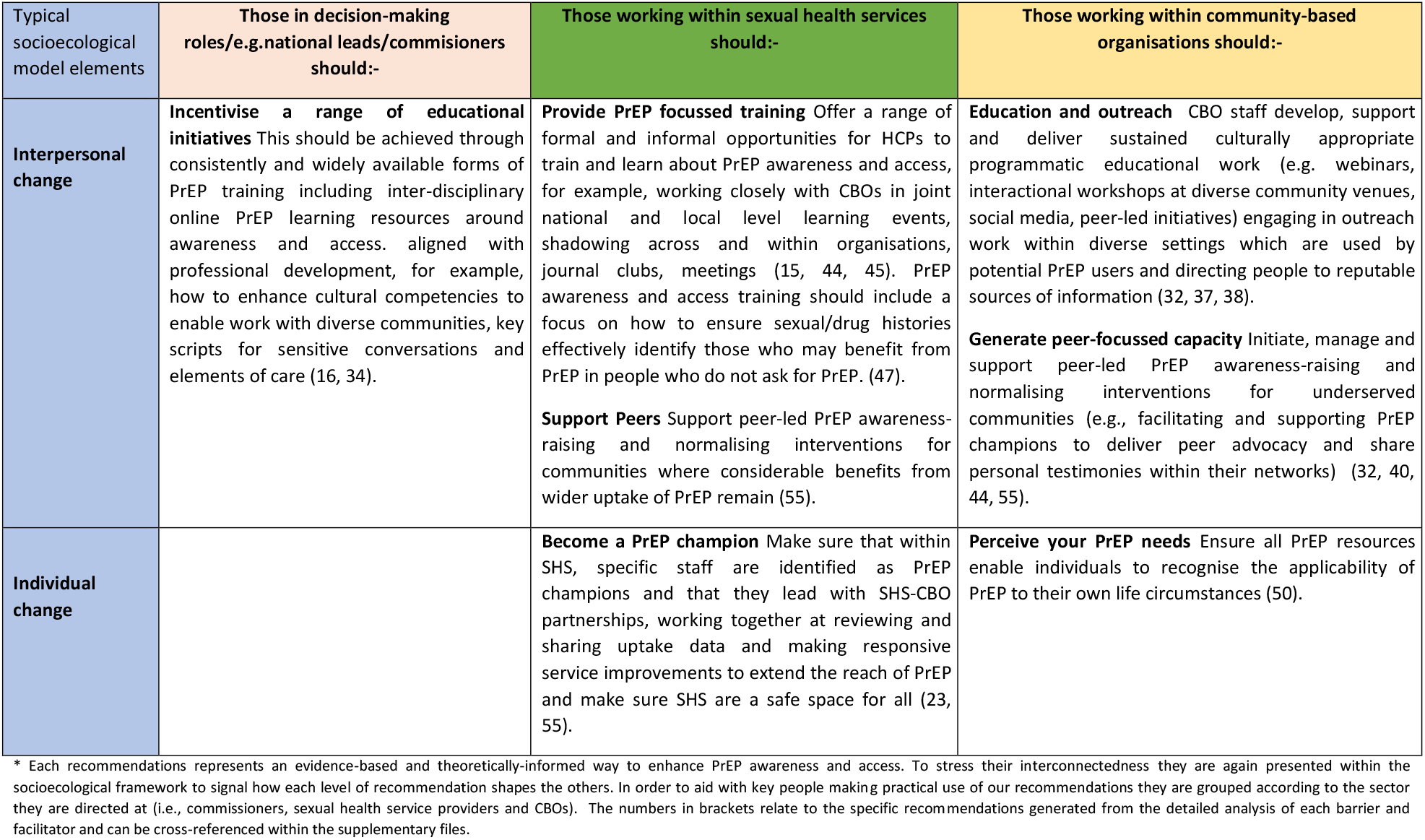
Recommendations for improving PrEP awareness and access at the microsical level.

At the macrosocial level, political will and momentum should be harnessed to drive the development and maintenance of networks of stakeholders across diverse health and social care organisations. These networks should include a broad range of statutory, and community-based organisations to facilitate exchange of skills, lessons-learned and data sharing. Program commissioners should forge effective partnerships across this network (e.g., partnerships between sexual health and addiction services) and encourage joint posts, initiatives and regular meetings. CBOs working with a broad range of communities should ensure initiatives are in place to support PrEP users to take up policy development roles, ensuring those affected by PrEP are well placed to help shape all key decisions. In relation to future generations, government should ensure that young people receive comprehensive relationship and sex education that incorporates PrEP and addresses the specific needs of particular communities (e.g., trans people, people of Black African ethnicity). At the mesosocial level, we recommend establishing monitoring systems that regularly appraise progress in relation to awareness and access. For those in major decision-making roles (e.g., government) such systems should appraise the effectiveness of the cross-sector networks and the partnerships within them; for sexual health services, monitoring systems should include measures of PrEP uptake and reach into communities who could further benefit from PrEP; for those in community organisations, assessment could include the added value of novel partnerships with organisations such as those which traditionally focus on mental health or poverty.

The recommendations at the mesosocial level also suggest that PrEP services are expanded to include provision in other settings e.g. primary care, or community outreach services Critically, across sexual health services and CBOs, effective innovations that increase awareness and access should be formally shared. This means there should be clear processes in place to disseminate any lessons learned about what works well, or what to avoid (e.g., transferring insights into harnessing peer influence between communities).

Both formal and informal opportunities should be provided for peer learning (e.g., shadowing, or the use of protocols and scripts to share good practice). We recommend that organisational cultures are enhanced to ensure that they enable access and that these cultures, once established are actively maintained (and monitored). The generation of such cultures should be driven by stakeholder engagement and the use of data and active monitoring. A system akin to the ‘Investors in people’ accreditation standards [28] could also be co-produced across the sector. This would benchmark services and assure all service-users that holistic and inclusive understandings of sexual health and wellbeing are central and valued within organisations.

We also recommend that diverse, multi-level, awareness-raising interventions should be established and sustained. These interventions should be co-created with the different communities with which they seek to engage. CBOs can play a particular role in making HIV and PrEP more salient in the lives and priorities of communities who could further benefit from PrEP (e.g. drug using communities). Furthermore, coproduction approaches across the sector (community representatives, CBO staff and HCPs) should also provide tools for sexual health services and other providers in the form of culturally appropriate scripts and wider resources. We also recommend that CBOs are encouraged to instigate, develop and evaluate novel outreach (e.g., for Black African communities or trans communities).

At the microsocial level many of the recommendations detailed above should be operationalised through PrEP-focussed training. This should include sector-wide provision of peer-learning and support for diverse professionals.

Complementing this work with professionals, we also recommend focussed activities to normalise PrEP and increase awareness within the diverse communities who could benefit from PrEP. This could involve CBOs facilitating and supporting peer advocacy within networks of minoritized groups who are under-served in relation to PrEP. At the individual level, staff from sexual health services or CBOs should become ‘PrEP champions’ with a duty to maintain and demonstrate PrEP-positive organisational culture.

Finally, there is a need to ensure that all PrEP resources systematically enable individuals to recognise the applicability of PrEP to their particular life circumstances.

## DISCUSSION

### Main findings

This work is the first to address a significant evidence gap regarding exactly *how* to improve awareness of and access to PrEP. Awareness and access are fundamental to effective use of PrEP for HIV prevention as they lead to uptake and adherence. Our findings suggest that improved awareness and access needs concerted action across multiple, interdependent and overlapping macro-social, meso-social and micro-social levels involving both healthcare and community systems. It involves the co-ordination of multiple institutions, organisations, communities, teams and individual professionals, patients and the public. The simultaneous cascading of multiple and multi-levelled interdependent changes stands a far higher chance of eliciting sustainable change than any single isolated innovation.[29] Complex and adaptive systems such as health and social care are highly resistant to long term change. To achieve a tipping point within such systems, and to enable the desired emergent effects of genuine changes to awareness and access, targeted and multi-levelled interventions are required. Our analyses suggest where these recommended intervention are needed most and key content. There is increasing recognition of the need for this kind of ‘whole-system’ intervention within public health along with a growing awareness of the concomitant challenges of their implementation.[30] Our findings contribute to the field by specifying practical and accessible recommendations to change the whole system, using novel and auditable methods.

#### Findings in context

There has been a long-standing acknowledgement of the challenges of increasing awareness and access to PrEP.[31-34] The barriers and facilitators to PrEP awareness and access we found were multifaceted and systemic. Problems at the policy-level were also manifested at the organisational, community, interpersonal and individual levels. Political will and cross-sector activism were vital to kick-starting the national programme, but they did so against a background of long-standing and seemingly intractable social injustice (e.g., homophobia, structural racism, sex-related stigma). This tension between biomedical promise and innovation (i.e., ‘a pill to prevent HIV’), and the complexity of the social systems in which it must gain traction, sits at the heart of PrEP’s role in elimination of HIV transmission. This tension patterns many of our findings and addressing it is key to effective PrEP implementation.[3, 4] At the organisational level, the widespread lack of additional resource to fund the implementation of the PrEP programme, heightened anxieties about already overstretched sexual health services. Key facilitators included the re-organisation of existing resource (e.g., use of nurse prescribing) and the normalisation of PrEP interactions over time (getting over the main innovation hump). Fundamental barriers included a lack of diverse settings in which to access PrEP and services lacking cultural competency. The latter very effectively demonstrated the complex interplay across the macrosocial, mesosocial and microsocial levels and the overarching role of such macrosocial drivers such as historic lack of sexual health education at the policy level. We have previously noted that building cultural competency as a component of improving sexual health literacy is a requirement for sustainable and equitable implementation of sexual healthcare and prevention.[35] At the community level, our findings highlight how attempts to increase awareness and access to PrEP were influenced by long-standing health inequalities. There is a growing literature highlighting this as a key issue.[3, 36-39] In Scotland the decision not to include an early mass media intervention to promote awareness (to buffer services from a surge of people seeking PrEP) meant that existing inequalities were amplified. Prior to PrEP becoming freely available to those who could benefit, some people (e.g., some gay communities) already had the connections to be reached by community norms promoting PrEP, and others did not.[26] Together, the extent and variety of these findings suggest the need for future interventions to enhance PrEP awareness and access to be wide ranging, co-ordinated and multi-levelled.

This work moves the field forwards with evidence-based recommendations grounded within what has been learned from Scotland and detailing suggestions of what is likely to be useful elsewhere. Political will and momentum should be harnessed to maintain a network of stakeholders and drive and incentivise partnerships. In turn, in relation to awareness and access, organisations must put monitoring systems in place, to evaluate their on-going efforts, learn more about themselves, each other, and their interactions and actively share what they are learning. In addition, ambitious multi-levelled awareness raising interventions and approaches need to be co-created with the diverse communities they seek to engage, recognising the role of expertise from across communities and CBOs in addition to specialist clinical and academic knowledge. In parallel, co-created accreditation systems could help e.g. to assure potential service users of organisational cultural-competence.

### Strengths and limitations

To our knowledge this is the first study which systematically examines and explores findings from the initial stages of a nationally implemented PrEP programme. Our approach illustrates exactly how it is possible to go further than earlier conceptualisations of PrEP implementation, (e.g.,[11-13]) and use multi-perspectival qualitative data, and multiple analytic tools (e.g., TDF, BCTT, socioecological model) to generate evidence-based and theoretically informed recommendations to enhance PrEP awareness and access to PrEP.

Our systematic approach has given us confidence in our findings which would have been difficult to achieve had we generated our recommendations through other approaches commonly used in recommendation development, meta-synthesis of implementation studies, expert opinion alone, sole reliance on public and patient involvement, or interpreting barriers and facilitators and making ‘common-sense’ judgements about what needs to change.

Our systematic approach reiterates and specifies key lessons-learned from the Scottish roll out of PrEP but also details novel content via the application of our implementation science tools. Our recommendations are firstly determined by granular analyses of the qualitative implementation insights (e.g., akin to those delivered by typical thematic analyses). Secondly, our recommendations stem from our systematic theorisation (i.e., using the TDF) of the barriers and facilitators of prior attempts to deliver to awareness and access. Finally, capitalising on our use of theory, we have then been able to systematically generate recommendations for future awareness raising and access promoting activities using the BCW and the BCTT. All recommendations are auditable in terms of examining which intervention components they stemmed from (i.e., ‘intervention functions’, ‘BCTs’), how they were theorised (i.e., using the TDF) and which specific barriers and facilitators they relate to. Critically, all are drawn from our participant voices. Other strengths include our commitment to stakeholder engagement through the use of multi-perspectival work, expert opinion and wider stakeholder involvement.

Limitations include our reliance on qualitative data alone; our assessment of the relative importance to key barriers and facilitators was not triangulated with quantitative data collected across nationally representative samples of PrEP users, HCPs and CBOs. We acknowledge that data are from a single nation, which had a very specific model of PrEP delivery in which critically, PrEP was free for all service users. The study was conducted in the first two years of programme roll out and experiences will change as the programme matures. The lack of diversity in the demographic characteristics of participants, in particular a preponderance of white, cis-gendered GBMSM, also limits transferability to different settings, given the observed link between PrEP awareness and access and existing health inequalities.

#### Implications for policy and practice

This systematic generation of recommendations from the Scottish experience has most relevance within the UK, Europe and Australia but these recommendations will also be relevant in other countries with similar health care systems and communities affected by HIV. Our recommendations may be used as direct tools to structure focussed-service planning maximising PrEP awareness and access. For example, using relevant, national and local stakeholders, either the synthesised recommendations, or the more granular recommendations presented in the Supplementary files, could be used to structure discussion and plan implementation effectively, translating our recommendations to the local context.

However, as our work with the socioecological framework shows, a multi-layered and multi-agent approach is prerequisite to enhancing PrEP awareness and access. This is because reinforcing positive feedback loops (i.e., where harnessing change to one aspect within a system reinforces change elsewhere) are likely to be generated between and across the individual recommendations, leading to systems wide change.[29] Future research is needed to evaluate the full impact of the recommendations we present within this paper and others (Estcourt et al., in preparation; MacDonald et al., in preparation). Together they represent a highly complex intervention [40] amenable to both process and outcome evaluation through a natural experimental design.

The use of HIV medication to prevent HIV acquisition has been described as a “game-changer”. Our work provides detailed analysis, based in lived experience, on how this can be implemented more effectively in future. Each step towards better PrEP implementation will be a step towards making HIV transmission a rarer event, and preventing the costly consequences of HIV infection.

## Data Availability

No data from this qualitative data set is publicly available. Supplementary file B provides an auditable account of anonymised data analysis.

## Acknowledgements

We are very grateful to the users, patients and staff of sexual health services in all 14 Health Boards within Scotland, Drs Ruth Holman, Maggie Gurney, Nil Banerjee, Pauline McGough, Daniela Brawley, Kirsty Abu-Rajab, Hame Lata, Anne McLellan, Alison Currie, Sharon Cameron, Hilary MacPherson, Janice Irvine, Graham Leslie, Ciara Cunningham, Maggie Watts. We thank staff and users of HIV Scotland; Waverley Care (SX Project and African Health Project); THT Scotland; Hwupenyu Health and Wellbeing; and Scottish Trans Alliance. We thank Nathan Sparling, Jacqueline Gray and Robson Dodd for their contributions to the research process.

## Competing Interests statement

The investigators named have no financial interests that impact on their responsibilities towards the scientific value or potential publishing activities associated with the study. However, the team has other interests within the field including various roles relating to HIV and sexual health within Government (Steedman, Estcourt, Nandwani, Clutterbuck), policy generation (Steedman, Nandwani, Estcourt, Saunders, Young, Flowers, HIV Scotland), practice (Steedman, Estcourt, Nandwani, Clutterbuck, Saunders) and advocacy (Young, HIV Scotland). PF reports research grants from National Institute of Health Research UK, Chief Scientist Office of Scotland. RN reports research grants from National Institute of Health Research UK, Chief Scientist Office of Scotland and non-executive director membership of the Board of Public Health Scotland from April 2020. JF reports research grants from Chief Scientist Office of Scotland. CSE reports research grants from National Institute of Health Research UK, Chief Scientist Office of Scotland, Engineering and Physical Sciences Research Council, UK Clinical Research Collaboration, Health Protection Scotland, European Centres for Disease Control. JM, JS, IY, DC, NS, LM & JD report no competing interests.

## Funding Statement

This work was supported by Chief Scientist Office grant number [HIPS/17/47].

## Author Contributions

All authors contributed to the conception and design of the studies, interpretation of findings, revision of the manuscript and approved the final version. Specific additional contributions are as follows: PF was co-investigator and led the qualitative design and behavioural science. PF wrote the initial draft of the manuscript. JM led the study day to day and undertook all research activities including data collection and analysis under the supervision of PF and CSE. JS, RN, DC, NS and CSE provided expert clinical interpretation. IY and JF contributed to data collection and analysis. JD led the ethical approval application. CSE was principal investigator and involved in all stages of the research.

## Ethical considerations

The study received ethical approval from the Glasgow Caledonian University Research Ethics Committee (REC) (HLS/NCH/17/037, HLS/NCH/17/038, HLS/NCH/17/044) and the South East of Scotland NHS REC (18/SS/0075, R&D GN18HS368).

## Notes

### Funding Statement

This study was funded by Chief Scientist Office grant number [HIPS/17/47].

### Author Declarations

The Glasgow Caledonian University Research Ethics Committee (REC) (HLS/NCH/17/037, HLS/NCH/17/038, HLS/NCH/17/044) and the South East of Scotland NHS REC (18/SS/0075, R&D GN18HS368) gave this study ethical approval.

## References

1 Fonner VA, Dalglish SL, Kennedy CE, et al. Effectiveness and safety of oral HIV preexposure prophylaxis for all populations. AIDS. 2016;30(12):1973–83. doi:10.1097/QAD.0000000000001145 [published Online First: 13 July 2016].

2 UNAIDS. Oral pre-exposure Prophylaxis: Putting a new choice in context. Joint United Nations Programme on HIV/AIDS (UNAIDS). 2015. https://www.who.int/hiv/pub/prep/who-unaids-prep-2015.pdf [accessed 12/03/22].

3 Ezennia O, Geter A, Smith DK. The PrEP care continuum and black men who have sex with men: a scoping review of published data on awareness, uptake, adherence, and retention in PrEP care. AIDS Behav. 2019;23(10):2654–73. doi:10.1007/s10461-019-02641-2 [published Online First: 28 August 2019].

4 Mayer KH, Allan-Blitz LT. PrEP 1.0 and beyond: optimizing a biobehavioral intervention. J Acquir Immune Defic Syndr. 2019;82(2):S113–S117. doi:10.1097/QAI.0000000000002169 [published Online First: 29 October 2019].

5 UNAIDS. Fast-track. Ending the AIDS epidemic by 2030. Geneva, Switzerland: Joint United Nations Programme on HIV/AIDS (UNAIDS). 2014. https://files.unaids.org/en/media/unaids/contentassets/documents/unaidspublication/2014/20140925_Fast_Track_Brochure.pdf [accessed 12/03/22].

6 ECDC. Country case studies - HIV pre-exposure Prophylaxis in the EU/EEA and the UK: implementation, standards and monitoring. European Centre for Disease Prevention and Control (ECDC). 2020. https://www.ecdc.europa.eu/sites/default/files/documents/Country-case-studies-PrEP.pdf [accessed 12/03/22]..

7 Gilbert L and Walker L. Treading the path of least resistance: HIV/AIDS and social inequalities—a South African case study. Soc Sci Med. 2002;54(7):1093–110. doi:10.1016/s0277-9536(01)00083-1 [published Online First: 18 February 2002].

8 Rhodes T, Singer M, Bourgois P, et al. The social structural production of HIV risk among injecting drug users. Soc Sci Med. 2005;61(5):1026–44. doi:10.1016/j.socscimed.2004.12.024 [published Online First: 19 March 2005].

9 UNAIDS. Confronting inequalities: Lessons for pandemic responses from 40 years of AIDS. Joint United Nations Programme on HIV/AIDS (UNAIDS) Global AIDS update. 2021. https://www.unaids.org/sites/default/files/media_asset/2021-global-aids-update_en.pdf [accessed 12/03/22].

10 Zablotska IB, Baeten JM, Phanuphak N, et al. Getting pre-exposure prophylaxis (PrEP) to the people: opportunities, challenges and examples of successful health service models of PrEP implementation. Sex Health. 2018;15(6):481–4. doi:10.1071/SH18182 [published Online First: 1 January 2018].

11 Liu, A., Colfax, G., Cohen, S., et al. (eds.) The spectrum of engagement in HIV prevention: proposal for a PrEP cascade. In 7th International conference on HIV treatment and prevention adherence, Florida: Miami Beach; 2012. p. 3–5.

12 Kelley CF, Kahle E, Siegler A, et al. Applying a PrEP continuum of care for men who have sex with men in Atlanta, Georgia. Clin Infect Dis. 2015;61(10):1590–7. doi:10.1093/cid/civ664 [published Online First: 13 August 2015].

13 Nunn AS, Brinkley-Rubinstein L, Oldenburg CE, et al. Defining the HIV pre-exposure prophylaxis care continuum. AIDS. 2017;31(5):731–34. doi:10.1097/QAD.0000000000001385 [published Online First: 1 Mach 2017].

14 Pinto RM, Berringer KR, Melendez R, et al. Improving PrEP implementation through multilevel interventions: a synthesis of the literature. AIDS Behav. 2018;22(11):3681–91. doi:10.1007/s10461-018-2184-4 [published Online First: 5 June 2018].

15 Zhang L, Peng P, Wu Y, et al. Modelling the epidemiological impact and cost-effectiveness of PrEP for HIV transmission in MSM in China. AIDS Behav. 2019;23(2):523–33. doi:10.1007/s10461-018-2205-3 [published Online First: 3 July 2018].

16 Koechlin FM, Fonner VA, Dalglish SL, et al. Values and preferences on the use of oral preexposure prophylaxis (PrEP) for HIV prevention among multiple populations: a systematic review of the literature. AIDS Behav. 2017;21(5):1325–35. doi:10.1007/s10461-016-1627-z [published Online First: 29 November 2016].

17 Presseau J, McCleary N, Lorencatto F, et al. Action, actor, context, target, time (AACTT): a framework for specifying behaviour. Implementation Sci. 2019;14(1):1–3. doi:10.1186/s13012-019-0951-x [published Online First: 5 December 2019].

18 Whiteley D, Speakman E, Elliot L, et al. Developing a primary care-initiated hepatitis C treatment pathway in Scotland: A qualitative study. Br J Gen Pract. Published Online First: 21 March 2022. doi:10.3399/BJGP.2022.0044

19 Atkins L, Francis J, Islam R, et al. A guide to using the Theoretical Domains Framework of behaviour change to investigate implementation problems. Implementation Sci. 2017;12(1):1–8. doi:10.1186/s13012-017-0605-9

20 Michie S, Atkins L, West R. The Behaviour Change Wheel: A Guide to Designing Interventions. London: Silverback Publishing; 2014.

21 Michie S, Richardson M, Johnston M, et al. The behavior change technique taxonomy (v1) of 93 hierarchically clustered techniques: Building an international consensus for the reporting of behavior change interventions. Ann Behav Med, 2013;46(1):81–95. doi:10.1007/s12160-013-9486-6 [published Online First: 20 March 2013].

22 Bronfenbrenner U. The ecology of human development: Experiments by nature and design. Cambridge, Mass: Harvard University Press 1979.

23 Health Protection Scotland. Implementation of HIV PrEP in Scotland Second Year Report: An Official Statistics publication for Scotland (Experimental). Health Protection Scotland, NHS National Services Scotland. 2019. https://hpspubsrepo.blob.core.windows.net/hps-website/nss/2914/documents/1_2019-12-17-HIV-PrEP-Implementation-Summary.pdf [accessed 12/03/22].

24 Estcourt C, Yeung A, Nandwani R, et al. Population-level effectiveness of a national HIV preexposure prophylaxis programme in MSM. AIDS. 2021;35(4):665. doi:10.1097/QAD.0000000000002790

25 Grimshaw C, Estcourt CS, Nandwani R, et al. Implementation of a national HIV pre-exposure prophylaxis service is associated with changes in characteristics of people with newly diagnosed HIV: a retrospective cohort study. Sex Transm Infect. 2022;98(1):53-7. doi:http://dx.doi.org/10.1136/sextrans-2020-054732 [published Online First: 13 January 2021].

26 Young I. Anticipating policy, orienting services, celebrating provision: Reflecting on Scotland’s PrEP journey. In Bernays S, Bourne A, Kippax S, et al eds. Remaking HIV Prevention in the 21st Century Cham: Springer 2021:59–72. https://link.springer.com/chapter/10.1007/978-3-030-69819-5_5 [accessed 12/03/22].

27 Braun V, Clarke V. Using thematic analysis in psychology. Qual Res Psychol. 2006;3(2):77–101. http://eprints.uwe.ac.uk/11735 [accessed 12/03/22].

28 Alberga T, Tyson S, Parsons D. An evaluation of the Investors in People Standard. Human Resource Management Journal. 1997;7(2):47–60. doi:https://doi.org/10.1111/j.1748-8583.1997.tb00281.x

29 Rutter H, Savona N, Glonti K, et al. The need for a complex systems model of evidence for public health. The Lancet. 2017;390(10112):2602–4. doi:10.1016/S0140-6736(17)31267-9 [published Online First 13 June 2017].

30 Jebb SA, Finegood DT, Diez Roux A, et al. Systems-based approaches in public health: where next?. The Academy of Medical Sciences. 2021. https://acmedsci.ac.uk/file-download/73812388 [accessed 12/03/22].

31 Hankins CA. Untangling the cost–effectiveness knot: who is oral antiretroviral HIV pre-exposure prophylaxis really for?. Expert Rev Pharmacoecon Outcomes Res. 2014;14(2):167–70. doi:10.1586/14737167.2014.887447 [published Online First: 19 February 2014].

32 Lachowsky NJ, Tattersall TL, Sereda P, et al. Community awareness of, use of and attitudes towards HIV pre-exposure prophylaxis (PrEP) among men who have sex with men in Vancouver, Canada: preparing health promotion for a publicly funded PrEP program. Sex Health. 2019;16(2):180–6. doi:https://doi.org/10.1071/SH18115

33 Newman R, Katchi T, Karass M, et al. Enhancing HIV pre-exposure prophylaxis practices via an educational intervention. Am J Ther. 2019;26(4):e462–8. doi:10.1097/MJT.0000000000000773

34 Young I, Valiotis G. Strategies to support HIV literacy in the roll-out of pre-exposure prophylaxis in Scotland: findings from qualitative research with clinical and community practitioners. BMJ open. 2020;10(4):e033849. doi:10.1136/bmjopen-2019-033849

35 McDaid L, Flowers P, Ferlatte O, et al. Sexual health literacy among gay, bisexual and other men who have sex with men: a conceptual framework for future research. Cult Health Sex. 2021;23(2):207–23. doi:10.1080/13691058.2019.1700307 [published Online First: 2 March 2020].

36 Philbin MM, Parker CM, Parker RG, et al. The promise of pre-exposure prophylaxis for Black men who have sex with men: an ecological approach to attitudes, beliefs, and barriers. AIDS Patient Care STDS. 2016;30(6):282–90. doi:10.1089/apc.2016.0037 [published Online First: 24 May 2016].

37 Rowniak S, Ong-Flaherty C, Selix N, et al. Attitudes, beliefs, and barriers to PrEP among trans men. AIDS Educ Prev. 2017;29(4):302–14. doi:10.1521/aeap.2017.29.4.302

38 Witzel TC, Nutland W, Bourne A. What are the motivations and barriers to pre-exposure prophylaxis (PrEP) use among black men who have sex with men aged 18-45 in London? results from a qualitative study. Sex Transm Infect. 2019;95:262–6. doi:10.1136/sextrans-2018-053773

39 Nakasone SE, Young I, Estcourt CS, et al. Risk perception, safer sex practices and PrEP enthusiasm: barriers and facilitators to oral HIV pre-exposure prophylaxis in Black African and Black Caribbean women in the UK. Sex Transm Infect. 2020 Aug 1;96(5):349–54. doi:10.1136/sextrans-2020-054457 [published Online First: 12 June 2020].

40 Skivington K, Matthews L, et al. A new framework for developing and evaluating complex interventions: update of Medical Research Council guidance. BMJ. 2021;374:n2061. doi:https://doi.org/10.1136/bmj.n2061

