## Supplementary Table A for "How can we enhance HIV Pre Exposure Prophylaxis (PrEP) awareness and access?: Recommendation development from process evaluation of a national PrEP programme using implementation science tools"

**Where should we focus future interventions to increase awareness of and access to PrEP?**

Figure 2 The selection of priority areas for future interventions to increase awareness and access to PrEP.

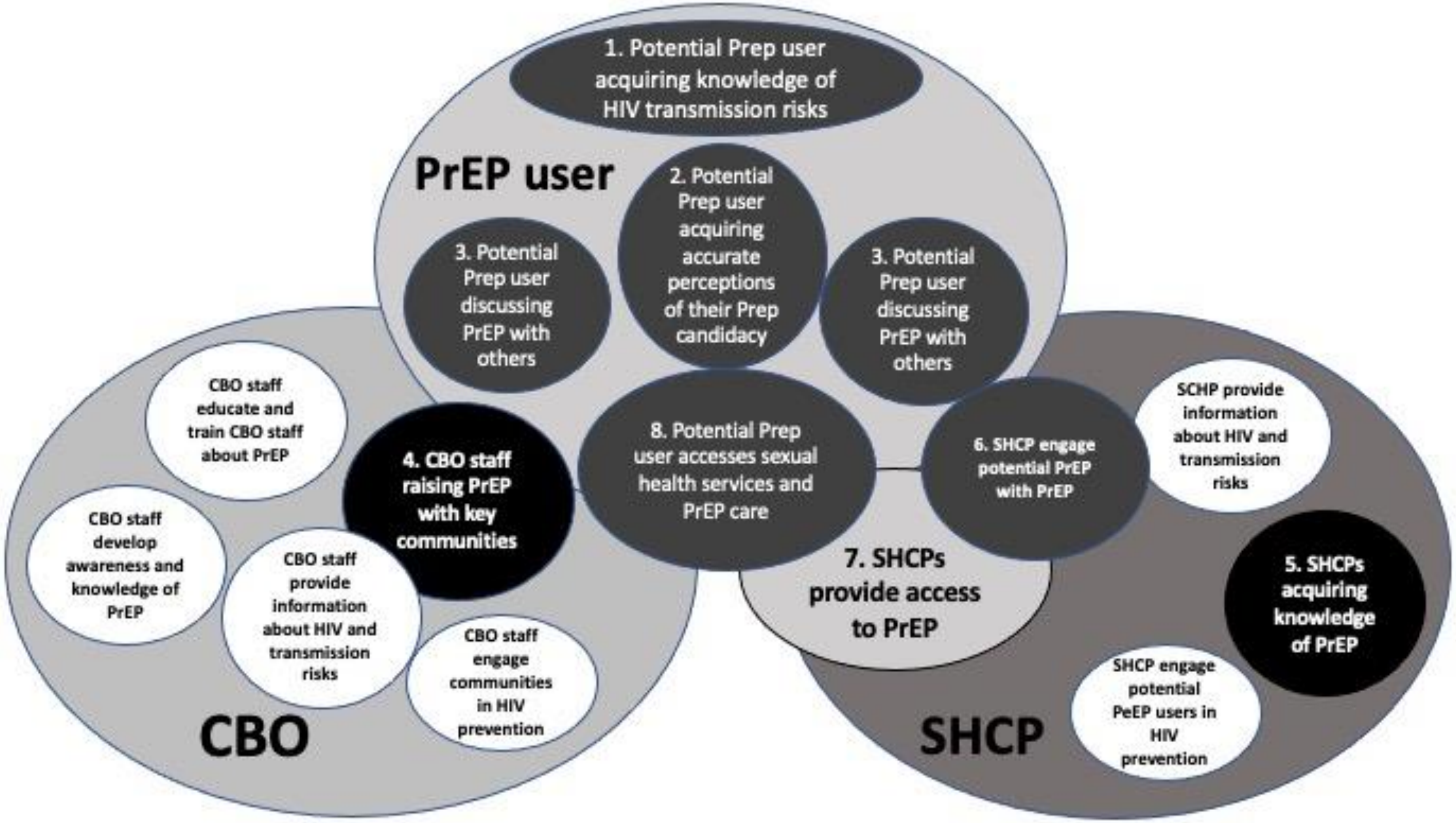

Legend: Text in black ovals depict priority areas for future intervention, text in white ovals shows areas that were not seen as priority areas for action. Selected areas were: (1) Potential PrEP users acquiring knowledge of HIV and its transmission risks in addition to acquiring knowledge of PrEP itself; (2) Potential PrEP users acquiring accurate perceptions of their PrEP candidacy; (3) PrEP users discussing PrEP with others; (4) CBO staff raising issues of PrEP with key communities; (5) HCPs acquiring knowledge of PrEP; (6) HCPs engage potential PrEP users with PrEP; (7) Sexual health services provide access to PrEP; and (8) Potential PrEP users access sexual health services and PrEP care therein.
