## Supplementary Table B for "How can we enhance HIV Pre Exposure Prophylaxis (PrEP) awareness and access?: Recommendation development from process evaluation of a national PrEP programme using implementation science tools"

**Supplementary files: Matrices of the intervention development analyses used to theorise key barriers and facilitators to develop evidence-based and theoretically informed recommendations for future interventions to improve PrEP awareness and access**

### Priority area 1: Potential PrEP user acquiring knowledge of HIV transmission risks

| Priority areas for intervention | Barriers | Facilitators | Indicative quotes | TDF domains | Intervention functions | Potential BCTs | Agreed final recommendations for those considering implementing PrEP at scale- post APEASE<br><br>Green text – consensus about inclusion<br>Brown text – some amendments made<br>Red text – rejected and not taken further | Framing within the Socio-ecological model |
|---|---|---|---|---|---|---|---|---|
| **Potential PrEP user acquiring knowledge of HIV transmission risks** | Potential PrEP users find it difficult to acquire knowledge of HIV transmission risks because of the absence of a marketing campaign to promote PrEP. | Potential PrEP users find it easy to acquire knowledge of HIV transmission risks because they have access to PrEP coverage within the gay press, on social media and dating apps and hook-up sites, and through informal peer education. | **Example 1**<br>*"The only way the closet queen married guy is going to find out about PrEP, or the one who's not going out on the scene any more or not using social apps, it's through media, they're not going to hear about it otherwise. But they were reluctant, they've always been reluctant to do any kind of national campaign."* (CBO staff working with GBMSM)<br><br>**Example 2**<br>*"I use some gay dating apps and a lot of people mentioned on their profile that they use PrEP, and at some point I kind of asked around and I found out that this is something that* | Knowledge<br><br>Environmental context and resources | Education<br><br>Environmental restructuring<br><br>Enablement | 5.1 Information about health consequences<br><br>5.3 Information about social and environmental consequences<br><br>5.6 Information about emotional consequences<br><br>9.1 Credible source<br><br>12.2 Restructure the social environment | 31. Ensuring clinical capacity was available and coinciding with PrEP roll-out, public health agencies, health authorities, and others with a remit for sexual health promotion should commission a mass media/social marketing campaign aimed at reaching all those who may benefit from PrEP. This could be fronted by culturally appropriate opinion leaders and would aim to share news of recent advancements within the HIV field (e.g. U=U, PrEP) and inform about the economic and wider benefits and value of PrEP for the healthcare system, communities, and individuals (5.1, 5.3, 5.6, 9.1)<br><br>32. Ensure that all potential PrEP users from a variety of | **31. Community change**<br><br>32.**Community change** |

**Supplementary files: Matrices of the intervention development analyses used to theorise key barriers and facilitators to develop evidence-based and theoretically informed recommendations for future interventions to improve PrEP awareness and access**

| Priority areas for intervention | Barriers | Facilitators | Indicative quotes | TDF domains | Intervention functions | Potential BCTs | Agreed final recommendations for those considering implementing PrEP at scale- post APEASE<br><br>Green text – consensus about inclusion<br>Brown text – some amendments made<br>Red text – rejected and not taken further | Framing within the Socio-ecological model |
|---|---|---|---|---|---|---|---|---|
| | | | *you could take to prevent you from getting HIV."* (PrEP user) | | | | communities have access to a range of PrEP-positive coverage, for example, within culturally relevant press and other media, in relevant settings (e.g., community centres, faith-based, trans-wellbeing venues, or sex-on-premises venues on targeted social media, via message blasts on dating apps and hook-up sites, and through informal peer education (12.2). | complemented by **Interpersonal change** |
| | Potential PrEP users find it difficult to acquire knowledge of HIV transmission risks because some other HCPs (e.g. GPs) have limited knowledge about or are | Potential PrEP users find it easy to acquire knowledge of HIV transmission risks because they attend SHS where SHCPs discuss HIV risks and prevention strategies and go through and provide a nationally- | **Example 1**<br>*"My own GP at the time knew nothing about PrEP, absolutely nothing at all. So I kind of just gave up on that avenue and realised that the clinic was the best place to talk about it."* (PrEP user)<br><br>**Example 2**<br>*"There was lots of papers there, I'm going to say leaflets, so basically just outlining the* | Knowledge<br><br>Professional role and identity<br><br>Environmental context and resources | Education<br><br>Persuasion<br><br>Modelling<br><br>Environmental restructuring<br><br>Training | 13.2 Framing /reframing<br><br>5.1 Information about health consequences<br><br>5.3 Information about social and environmental consequences | 33. If possible partnership work between CBOs and SHS should focus on educating other HCPs, such as GPs (e.g. during their initial training, CPD), provide information on key sexual health topics, including HIV risks, epidemiology, physical and psychosocial effects, and advances in treatment and prevention (e.g. U+U, PrEP) (5.1, 5.3, 5.6), and advice (e.g. clear statements and nuanced examples) regarding the eligibility | **33. Organisational change** |

**Supplementary files: Matrices of the intervention development analyses used to theorise key barriers and facilitators to develop evidence-based and theoretically informed recommendations for future interventions to improve PrEP awareness and access**

| Priority areas for intervention | Barriers | Facilitators | Indicative quotes | TDF domains | Intervention functions | Potential BCTs | Agreed final recommendations for those considering implementing PrEP at scale- post APEASE<br><br>Green text – consensus about inclusion<br>Brown text – some amendments made<br>Red text – rejected and not taken further | Framing within the Socio-ecological model |
|---|---|---|---|---|---|---|---|---|
| | unlikely to discuss sexual health issues. | developed patient information booklet(s). | *instructions on what it does, how often you have to take it, what the side-effects are…we went through all this literature together."* (PrEP user) | | | 5.6 Information about emotional consequences<br><br>4.1 Instruction on how to perform the behaviour<br><br>13.3 Incompatible beliefs<br><br>6.1 Demonstration of the behaviour<br><br>7.1 Prompts/cues<br><br>4.1 Instruction on how to perform the behaviour | criteria (4.1), and model ways for them to proactively and routinely discuss sexual health issues with clients (6.1).<br><br>34. Partnership work between CBOs, SHS and those involved in wider health services should develop guidance on key questions to ask when taking a sexual/ drug history, open-ended questions, and a tool to aid determination of risk based on answers (4.1, 6.1, 7.1).<br>35. Develop resources that should ideally be co-produced by a range of diverse organisations and the communities who will use them. These should promote accurate and consistent information-giving by developing a range of resources (e.g. fact sheet, PrEP provider pocket guide, national patient information booklets, 'how to' scripts) to help SHCPs and other | **34.Interpersonal change**<br><br>**35. Community change and interpersonal change** |

**Supplementary files: Matrices of the intervention development analyses used to theorise key barriers and facilitators to develop evidence-based and theoretically informed recommendations for future interventions to improve PrEP awareness and access**

| Priority areas for intervention | Barriers | Facilitators | Indicative quotes | TDF domains | Intervention functions | Potential BCTs | Agreed final recommendations for those considering implementing PrEP at scale- post APEASE<br><br>Green text – consensus about inclusion<br>Brown text – some amendments made<br>Red text – rejected and not taken further | Framing within the Socio-ecological model |
|---|---|---|---|---|---|---|---|---|
| | | | | | | | HCPs introduce PrEP and structure PrEP conversations (7.1, 4.1) and which clients can take away for further reading on HIV and PrEP (5.1, 5.3, 5.6). | |
| | Potential PrEP users find it difficult to acquire knowledge of HIV transmission risks because they struggle to engage with the nationally-developed patient information booklets. | Potential PrEP users find it easy to acquire knowledge of HIV transmission risks because of SHS and CBO outreach initiatives. | **Example 1**<br>*"I mean, the best will in the world, nobody takes written information any more. We give hundreds of the PrEP leaflets out every time but most are left abandoned or ditched as soon as you're out the building."* (SHCP)<br><br>**Example 2**<br>*"I had given them the papers to read about PrEP before, which means the majority of them did not even bother to read. They just looked at it and said, I don't know what she's talking about, this PrEP, what is it. So this is where I'm trying to talk, to tell you, we have* | Knowledge<br><br>Environmental context and resources | Education<br><br>Environmental restructuring<br><br>Enablement | 5.1 Information about health consequences<br><br>5.3 Information about social and environmental consequences<br><br>5.6 Information about emotional consequences<br><br>12.1 Restructure the physical environment<br><br>9.1 Credible source | 36. Ensure that national patient information booklets aimed at educating potential PrEP users (and PrEP users) about HIV and PrEP are pitched at different levels to account for variations in HIV and PrEP literacy and individual preferences on how much information they receive (i.e. develop one booklet that covers the basics and another booklet that provides more comprehensive information) (5.1, 5.3, 5,6, 12.1).<br><br>37. SHCPs and CBO staff should direct clients to alternative reputable information sources (9.1) as well as providing national patient information booklets (e.g. | **36. Community change**<br><br>**37. Interpersonal change** |

**Supplementary files: Matrices of the intervention development analyses used to theorise key barriers and facilitators to develop evidence-based and theoretically informed recommendations for future interventions to improve PrEP awareness and access**

| Priority areas for intervention | Barriers | Facilitators | Indicative quotes | TDF domains | Intervention functions | Potential BCTs | Agreed final recommendations for those considering implementing PrEP at scale- post APEASE<br><br>Green text – consensus about inclusion<br>Brown text – some amendments made<br>Red text – rejected and not taken further | Framing within the Socio-ecological model |
|---|---|---|---|---|---|---|---|---|
| | | | *that oral kind of upbringing, where information is given to us orally. Therefore, our reading tends to be not so high on there. I think workshops would do the work. I mean, workshops where you actually go and share that information with someone…so then, it gets into their heads."* (CBO staff working with Black African communities) | | | 3.1 Social support (unspecified) | signpost to SHS, CBO, and HIV/PrEP activists' websites and social media) (3.1).<br><br>38. SHS and CBO should do outreach work to inform potential PrEP users about HIV and PrEP (e.g. black African community venues, sex-on-premises venues, trans community events, pop-up testing sites, interactional workshops at diverse community venues) (5.1, 5.3, 5.6, 12.1). | **38. Community change and embedded interpersonal change** |
| | Potential PrEP users find it difficult to acquire knowledge of HIV transmission risks because there is little advertising of PrEP in health | Potential PrEP users find it easy to acquire knowledge of HIV transmission risks because they receive a comprehensive sex education. | **Example 1**<br>*"In my doctors' surgery there is mention of things like LGBT issues and things like that but there is nothing on PrEP there."* (PrEP user)<br><br>**Example 2**<br>*"I think it [PrEP] should be talked about in schools. I don't know how much it is… but I think the more education we* | Environmental context and resources<br><br>Knowledge | Environmental restructuring<br><br>Education | 12.1 Restructure the physical environment<br><br>12.2 Restructure the social environment<br><br>5.1 Information about health consequences | 39. Develop a range of resources co-produced by SHS, CBO staff, and community representatives to promote PrEP (e.g. flyers, posters, national patient information booklets, short videos) and distribute these for display in non-sexual health-specific health services, such as reproductive health clinics, GP surgeries, pharmacies, and hospitals (12.1). | **39. Community change** |

**Supplementary files: Matrices of the intervention development analyses used to theorise key barriers and facilitators to develop evidence-based and theoretically informed recommendations for future interventions to improve PrEP awareness and access**

| Priority areas for intervention | Barriers | Facilitators | Indicative quotes | TDF domains | Intervention functions | Potential BCTs | Agreed final recommendations for those considering implementing PrEP at scale- post APEASE<br><br>Green text – consensus about inclusion<br>Brown text – some amendments made<br>Red text – rejected and not taken further | Framing within the Socio-ecological model |
|---|---|---|---|---|---|---|---|---|
| | settings other than SHS. | | *get out there about this, the better. Because nothing like this was talked about when I was at school, at all. And it's cost lives, the long and the short of it, we've lost people due to ignorance, to lack of education."* (PrEP user) | | | 5.3 Information about social and environmental consequences<br><br>5.6 Information about emotional consequences | 10. Governments should make age-appropriate and comprehensive relationships and sex education compulsory for children and young people at all levels of schooling (12.2), including content on the health, social, and emotional consequences of HIV and PrEP (5.1, 5.3, 5.6, 12.2). | **10. Public Policy change** |
| | | Potential PrEP users find it easy to acquire knowledge of HIV transmission risks because they have important others (e.g. friends, sexual partners) living with HIV who discuss their condition and advancements in HIV treatment and prevention. | *"I was aware of people that I've known that have had HIV and have had it for quite a long time and been on treatment and been almost survivors or such. So I knew there was a quite a lot of advancement."* (PrEP user) | Social influences<br><br>Knowledge | Environmental restructuring<br><br>Enablement<br><br>Education | 12.2 Restructure the social environment<br><br>5.1 Information about health consequences<br><br>5.4 Information about social and environmental consequences<br><br>5.6 Information about | 40. Public health agencies and those who provide HIV treatments should consider working with relevant CBOs to build on the activism and peer influence seen among MSM (12.2) and consider peer-led PrEP awareness-raising and normalising interventions (5.1, 5.3, 5.6, 9.1) for other communities where there may be considerable benefits from wider uptake of PrEP. | **40. Public Policy, Community, and Interpersonal change** |

**Supplementary files: Matrices of the intervention development analyses used to theorise key barriers and facilitators to develop evidence-based and theoretically informed recommendations for future interventions to improve PrEP awareness and access**

| Priority areas for intervention | Barriers | Facilitators | Indicative quotes | TDF domains | Intervention functions | Potential BCTs | Agreed final recommendations for those considering implementing PrEP at scale- post APEASE<br><br>Green text – consensus about inclusion<br>Brown text – some amendments made<br>Red text – rejected and not taken further | Framing within the Socio-ecological model |
|---|---|---|---|---|---|---|---|---|
| | | | | | | emotional consequences<br><br>9.1 Credible source | | |

**Supplementary files: Matrices of the intervention development analyses used to theorise key barriers and facilitators to develop evidence-based and theoretically informed recommendations for future interventions to improve PrEP awareness and access**

### Priority area 2: Potential PrEP user acquiring accurate perceptions of their PrEP candidacy

| Priority areas for intervention | Barriers | Facilitators | Indicative quotes | TDF domains | Intervention functions | Potential BCTs | Agreed final recommendations for those considering implementing PrEP at scale-post APEASE<br><br>Green text – consensus about inclusion<br>Brown text – some amendments made<br>Red text – rejected and not taken further | Framing within the Socio-ecological model |
|---|---|---|---|---|---|---|---|---|
| 1.Potential PrEP users acquire accurate perceptions of their PrEP candidacy. | Potential PrEP users find it difficult to acquire accurate perceptions of their PrEP candidacy because they believe that PrEP is for 'other' people (e.g. those who are at 'high-risk' for HIV). | | *"There was a part of me that thought, actually, this is an intervention that only people who are putting themselves at risk need, you know. Someone like me doesn't need it, because I'm not like that. But that's silly, of course, that was silly."* (PrEP user) | Knowledge<br><br>Beliefs about consequences | Education<br><br>Persuasion | 5.1 Information about health consequences<br><br>13.2 Framing /reframing | 41. Ensure PrEP information and communications (e.g. SHCP- and CBO staff-client interactions, national patient information booklets, SHS, CBO, and HIV/PrEP activists' websites and social media, marketing campaigns) educate potential PrEP users on the facts of HIV transmission (5.1), address PrEP-related stigma, for example, by adopting 'needs-based' terminology rather than focusing on 'risk', and provide advice (e.g. clear statements and nuanced examples) regarding the eligibility criteria (13.2). | **41. Community change** |
| | Potential PrEP users find it difficult to acquire accurate | Potential PrEP users find it easy to acquire accurate perceptions of | **Example 1**<br>*"I remember one of the persons who tried to access PrEP in [place],* | Knowledge<br><br>Beliefs about | Education<br><br>Persuasion | 5.1 Information about health consequences | 42. Promote accurate and consistent information-giving by educating other HCPs, such as GPs (e.g. during their initial | **42. Community change** |

**Supplementary files: Matrices of the intervention development analyses used to theorise key barriers and facilitators to develop evidence-based and theoretically informed recommendations for future interventions to improve PrEP awareness and access**

| Priority areas for intervention | Barriers | Facilitators | Indicative quotes | TDF domains | Intervention functions | Potential BCTs | Agreed final recommendations for those considering implementing PrEP at scale-post APEASE<br><br>Green text – consensus about inclusion<br>Brown text – some amendments made<br>Red text – rejected and not taken further | Framing within the Socio-ecological model |
|---|---|---|---|---|---|---|---|---|
| | perceptions of their PrEP candidacy because some other HCPs (e.g. GPs) have inadequate knowledge of PrEP and when / for whom it might be appropriate. | their PrEP candidacy because CBO staff provide expert advice. | *when it had just come out, they went to the GP and they were told, you know, it's not for you, it's for gay men, so the person was turned away, and when they phoned me, I was, like, okay, so what else do we need to do?"* (CBO staff working with Black African communities) | consequences<br><br>Environmental context and resources | Training<br><br>Enablement | 13.2 Framing /reframing<br><br>4.1 Instruction on how to perform the behaviour<br><br>7.1 Prompts/cues | training, CPD), on HIV risks and epidemiology and PrEP uses and efficacy (5.1), ensuring that training and resources to support PrEP discussions (e.g. fact sheet, PrEP provider pocket guide, national patient information booklets, 'how to' scripts) (4.1, 7.1) are explicit that PrEP is inclusive and relevant to all individuals with an identified need, not just GBMSM, and provide advice (e.g. clear statements and nuanced examples) regarding the eligibility criteria (13.2). | |
| | Potential PrEP users find it difficult to acquire accurate perceptions of their PrEP candidacy because some SHCPs are unsure how to | Potential PrEP users find it easy to acquire accurate perceptions of their PrEP candidacy because SHCPs are primed to assess and identify HIV risks | **Example 1**<br>*"Women who are at risk of HIV are probably pretty difficult to identify. I'd say, particularly people who are in a relationship, they're very difficult to identify. Particularly if they don't know that* | Environmental context and resources<br><br>Knowledge<br><br>Beliefs about | Enablement<br><br>Education<br><br>Training | 12.2 Restructuring the social environment<br><br>13.2 Framing/ reframing | 43. Create and uphold a service context that is harmonised with the goals of the PrEP programme by ensuring that PrEP information, training, education, and other communications directed at SHCPs are explicit that PrEP is inclusive and relevant to all individuals with an | **43. Organisational change** |

**Supplementary files: Matrices of the intervention development analyses used to theorise key barriers and facilitators to develop evidence-based and theoretically informed recommendations for future interventions to improve PrEP awareness and access**

| Priority areas for intervention | Barriers | Facilitators | Indicative quotes | TDF domains | Intervention functions | Potential BCTs | Agreed final recommendations for those considering implementing PrEP at scale- post APEASE<br><br>Green text – consensus about inclusion<br>Brown text – some amendments made<br>Red text – rejected and not taken further | Framing within the Socio-ecological model |
|---|---|---|---|---|---|---|---|---|
| | navigate the 'equivalent risk' eligibility criterion and fear that they might stigmatise or offend non-GBMSM clients by asking questions to assess PrEP candidacy. | among GBMSM clients (e.g. they expect GBMSM to be the main group accessing PrEP, view all GBMSM as potentially 'at-risk', have a clear sense of GBMSM HIV risks, and are used to talking to GBMSM about this). | *someone that they're having sex with has HIV. People from minority groups, they're quite difficult to identify. But we also don't know who we don't know about, at the moment. Because we have only had it for a year, so we haven't really got enough to data to know who are the people who we haven't identified."* (SHCP)<br><br>**Example 2**<br>*"We're all really well trained to know. If someone [an MSM] is telling you they're not using condoms for anal sex or they've had a few burst condoms, or they've split up with someone and they're having a bit* | consequences<br><br>Skills | | 5.1 Information about health consequences<br><br>4.1 Instruction on how to perform the behaviour<br><br>6.1 Demonstration of the behaviour<br><br>7.1 Prompts/cues<br><br>2.2 Feedback on behaviour<br><br>2.3 Self-monitoring of behaviour | identified need, not just GBMSM (12.2, 13.2).<br><br>44. SHS could consider outsourcing educational sessions for SHCPs to CBOs with expertise on the specific sexual health cultures of and HIV risks affecting Black Africans, trans people, and cisgendered women (5.1).<br><br>45. SHS could ask CBO staff who have high levels of cultural competency in delivering sexual health promotion interventions to Black Africans, trans people, and cisgendered women to share their tailored vocabularies and co-produce a stock of key phrases to enable SHCPs to sensitively probe clients when taking a sexual/drug history (4.1, 6.1, 7.1). | **44. Community change**<br><br>**45. Community change** |

**Supplementary files: Matrices of the intervention development analyses used to theorise key barriers and facilitators to develop evidence-based and theoretically informed recommendations for future interventions to improve PrEP awareness and access**

| Priority areas for intervention | Barriers | Facilitators | Indicative quotes | TDF domains | Intervention functions | Potential BCTs | Agreed final recommendations for those considering implementing PrEP at scale-post APEASE<br><br>Green text – consensus about inclusion<br>Brown text – some amendments made<br>Red text – rejected and not taken further | Framing within the Socio-ecological model |
|---|---|---|---|---|---|---|---|---|
| | | | *of a wild three months and they've been quite enjoying it, and this is something they think they might want to do for a bit longer. So, I think everyone's really confident at knowing straightway if someone [an MSM] would benefit from PrEP."* (SHCP) | | | 8.1 Behavioural practice/rehearsal<br><br>3.1 Social support (unspecified)<br><br>3.2 Social support (practical)<br><br>6.2 Social comparison | 46. Review and update the questions asked as part of a sexual/drug history on a regular basis to ensure they reflect the epidemiological evidence and any emerging new trends or behaviours which appear to enhance the risk of HIV and cascade any changes to all staff (4.1).<br><br>47. Ensure SHCPs maintain their knowledge of the HIV risks among different groups, including GBMSM, and skills in conducting culturally sensitive clinical risk assessments (e.g. ongoing CPD, clinical supervision) (5.1, 2.2, 2.3, 8.1).<br><br>48. Adopt a protocoled approach to PrEP that includes advice (e.g. clear statements and nuanced examples) regarding the eligibility criteria (4.1). | 46 **Organisational and interpersonal change**<br><br>**47. Organisational and interpersonal change**<br><br>**48. Organisational change** |

**Supplementary files: Matrices of the intervention development analyses used to theorise key barriers and facilitators to develop evidence-based and theoretically informed recommendations for future interventions to improve PrEP awareness and access**

| Priority areas for intervention | Barriers | Facilitators | Indicative quotes | TDF domains | Intervention functions | Potential BCTs | Agreed final recommendations for those considering implementing PrEP at scale-post APEASE<br><br>Green text – consensus about inclusion<br>Brown text – some amendments made<br>Red text – rejected and not taken further | Framing within the Socio-ecological model |
|---|---|---|---|---|---|---|---|---|
| | | | | | | | 49. Ensure a range of peer-support systems are in place (e.g. real-time/email support, team meetings, 'phone a friend', clinical network arrangements) to assist SHCPs in making complex eligibility decisions (12.2, 3.1, 3.2, 6.2). | **49. Organisational change and Public policy change** |
| | Potential PrEP users find it difficult to acquire accurate perceptions of their PrEP candidacy because PrEP information and communications tend to frame PrEP as primarily for GBMSM, to the exclusion of people from other HIV affected communities (e.g. Black Africans, | Potential PrEP users find it easy to acquire accurate perceptions of their PrEP candidacy because PrEP information and communications are explicit that PrEP is inclusive and relevant to all individuals with an identified need, not just GBMSM. | *"The way it was pitched to our communities, it's not for Africans, there is no clear messages that it's for Africans, it's around gay men, gay men, gay men, and that's been the messages constantly. So… it's not working for the African community, because the messages have to be strong and specific, the communities who can access PrEP. It's not just about gay men, and* | Environmental context and resources<br><br>Knowledge | Enablement<br><br>Education | 13.2 Framing /reframing<br><br>2.7 Feedback on outcome(s) of behaviour<br><br>5.1 Information about health consequences<br><br>5.3 Information about social and environmental consequences | 50. Ensure that all PrEP information and communications are explicit that PrEP is inclusive and relevant to all individuals (with an identified need) to enable groups other than GBMSM affected by HIV, such as Black Africans, trans people, and cisgendered women, to realise its applicability (13.2).<br><br>22. Partnership work between SHS, CBOs, and community representatives should co-produce tailored resources to raise awareness of HIV and PrEP | **50. Individual change**<br><br>**22. Community change** |

**Supplementary files: Matrices of the intervention development analyses used to theorise key barriers and facilitators to develop evidence-based and theoretically informed recommendations for future interventions to improve PrEP awareness and access**

| Priority areas for intervention | Barriers | Facilitators | Indicative quotes | TDF domains | Intervention functions | Potential BCTs | Agreed final recommendations for those considering implementing PrEP at scale-post APEASE<br><br>Green text – consensus about inclusion<br>Brown text – some amendments made<br>Red text – rejected and not taken further | Framing within the Socio-ecological model |
|---|---|---|---|---|---|---|---|---|
| | trans people, cisgendered women). | | *that's the way it's been. It needs to be a strong voice to say, it's for everybody."* (CBO staff working with Black African communities) | | | 5.6 Information about emotional consequences<br><br>12.1 Restructure the physical environment | and provide PrEP information (5.1, 5.3, 5.6) in the languages, tones, and formats most accessible by and acceptable to the intended audience, including advice (e.g. clear statements and nuanced examples) regarding the eligibility criteria (13.2, 2.7). Ensure these resources are disseminated and distributed through culturally appropriate means (12.1). | |
| | | Potential PrEP users find it easy to acquire accurate perceptions of their PrEP candidacy because SHCPs proactively contact clients who meet the eligibility criteria. | *"We would seek to identify any patients with a rectal bacterial STI and check if they've had a PrEP discussion and made a decision. If not, we would be contacting those patients and subsequently letting them know that our service provides PrEP and offering that service to them."* (SHCP) | Environmental context and resources<br><br>Knowledge | Enablement<br><br>Education | 3.1 Social support (unspecified)<br><br>5.1 Information about health consequences<br><br>2.7 Feedback on outcome(s) of behaviour | 51. At initial PrEP roll-out and routinely (e.g. quarterly) thereafter, SHCPs could run a report on the IT system to identify clients who (likely) meet the eligibility criteria but have not had a PrEP discussion and attempt to make contact via email, SMS, or phone to inform them about the health benefits of PrEP (5.1), its availability at the SHS (3.1), and their potential | **51 Organisational and interpersonal change** |

**Supplementary files: Matrices of the intervention development analyses used to theorise key barriers and facilitators to develop evidence-based and theoretically informed recommendations for future interventions to improve PrEP awareness and access**

| Priority areas for intervention | Barriers | Facilitators | Indicative quotes | TDF domains | Intervention functions | Potential BCTs | Agreed final recommendations for those considering implementing PrEP at scale-post APEASE<br><br>Green text – consensus about inclusion<br>Brown text – some amendments made<br>Red text – rejected and not taken further | Framing within the Socio-ecological model |
|---|---|---|---|---|---|---|---|---|
| | | | | | | 12.2 Restructure the social environment | eligibility (2.7) and offer a rapid appointment (12.2). | |

**Supplementary files: Matrices of the intervention development analyses used to theorise key barriers and facilitators to develop evidence-based and theoretically informed recommendations for future interventions to improve PrEP awareness and access**

**Priority area 3: Potential PrEP user discussing PrEP with others**

| Priority areas for intervention | Barriers | Facilitators | Indicative quotes | TDF domains | Intervention functions | Potential BCTs | Agreed final recommendations for those considering implementing PrEP at scale- post APEASE<br><br>Green text – consensus about inclusion<br>Brown text – some amendments made<br>Red text – rejected and not taken further | Framing within the Socio-ecological model |
|---|---|---|---|---|---|---|---|---|
| (Potential) PrEP users discussing PrEP with others. | (Potential) PrEP users find it difficult discussing PrEP with others because of stigmas around sex and sexual health in society generally. | (Potential) PrEP users find it easy discussing PrEP with others because SHCPs and CBO staff encourage them to talk about PrEP and support them to have PrEP conversations with important others | **Example 1**<br>*"As a society, we've created …almost, an approach to sex being quite, oh it's a bit of titillation, but it's wrong. Instead of looking at it as something we can do as part of the way we live, that is enjoyable. And it gives us a lot of pleasure, and it can help our mental health, you know, it can deal with anxiety, and is just good fun. So, we don't do that. And that makes it really difficult to have conversations with your sexual partners, because you're a little bit embarrassed, and a little* | Environmental context and resources<br><br>Social influences | Environmental restructuring<br><br>Enablement<br><br>Persuasion<br><br>Education | 12.2 Restructure the social environment<br><br>13.2 Framing/ reframing<br><br>1.2 Problem solving<br><br>5.1 Information about health consequences<br><br>5.3 Information about social and environmental consequences<br><br>5.6 Information about emotional consequences | 10. Governments should make age-appropriate and comprehensive relationships and sex education compulsory for children and young people at all levels of schooling, with fact-oriented and non-judgemental content that addresses the sexual health, social, and cultural needs of LGBTQ+ and Black African communities (12.2), incorporating issues such as PrEP as HIV prevention (5.1).<br><br>52. CBO staff and other HCPs (i.e. non-SHCPs), such as GPs, must address cultural stigmas and normalise talking about sex and sexual health-related issues, including HIV and PrEP, by engaging clients and the wider communities that they serve in | **10. Public Policy Change**<br><br>**52.Community change** |

**Supplementary files: Matrices of the intervention development analyses used to theorise key barriers and facilitators to develop evidence-based and theoretically informed recommendations for future interventions to improve PrEP awareness and access**

| Priority areas for intervention | Barriers | Facilitators | Indicative quotes | TDF domains | Intervention functions | Potential BCTs | Agreed final recommendations for those considering implementing PrEP at scale- post APEASE<br><br>Green text – consensus about inclusion<br>Brown text – some amendments made<br>Red text – rejected and not taken further | Framing within the Socio-ecological model |
|---|---|---|---|---|---|---|---|---|
| | | | *bit awkward, and a little bit, so I'm on PrEP, so it means we can have bareback sex, or condom-less sex, or whatever way you want to describe it."* (CBO staff working with MSM)<br><br>**Example 2**<br>*"I do actually say, look, you know, you make someone else have that conversation and bring up PrEP. Do you know about PrEP, you know, or if they've only been on PrEP a month, remember and say that you still might be in a window period, you might still be in an HIV window period. Yes, I think I try and encourage them to talk about PrEP."* (SHCP) | | | 9.1 Credible source<br><br>1.2 Problem Solving | topics of this nature (e.g. via discussions in everyday contexts / routine consultations, interactional workshops at diverse community venues, outreach work) (12.2).<br><br>53. Frame sex and sexual health as integral rather than peripheral to overall health and wellbeing, for example, during SHCP-, other HCP and CBO staff-client interactions, on SHS, CBO, and HIV/PrEP activists' websites and social media and posters in SHS, CBO, and non-sexual health service settings, and via sex and relationships education enhanced with cross-sector collaboration (13.2, 5.1, 5.3, 5.6).<br><br>54. SHCPs and CBO staff should encourage clients to discuss PrEP with important others by informing them of the important health, social, and emotional | 53. Organisational change, Community change, individual change<br><br>54. Organisational change, community |

**Supplementary files: Matrices of the intervention development analyses used to theorise key barriers and facilitators to develop evidence-based and theoretically informed recommendations for future interventions to improve PrEP awareness and access**

| Priority areas for intervention | Barriers | Facilitators | Indicative quotes | TDF domains | Intervention functions | Potential BCTs | Agreed final recommendations for those considering implementing PrEP at scale- post APEASE<br><br>Green text – consensus about inclusion<br>Brown text – some amendments made<br>Red text – rejected and not taken further | Framing within the Socio-ecological model |
|---|---|---|---|---|---|---|---|---|
| | | | | | | | benefits of doing so (e.g. increase awareness and uptake of PrEP, reduce PrEP-related stigma) (5.1, 5.3, 5.6, 9.1) and help to facilitate PrEP conversations by asking clients to identify potential barriers to talking about PrEP and selecting strategies to overcome these (1.2).<br><br>55. SHCPs and CBO staff could find ways of engaging and supporting PrEP champions from diverse communities to share their expertise and experiences with a wide audience of potential PrEP users (e.g. record a testimonial) (9.1). | change, Individual change |

**Supplementary files: Matrices of the intervention development analyses used to theorise key barriers and facilitators to develop evidence-based and theoretically informed recommendations for future interventions to improve PrEP awareness and access**

| Priority areas for intervention | Barriers | Facilitators | Indicative quotes | TDF domains | Intervention functions | Potential BCTs | Agreed final recommendations for those considering implementing PrEP at scale- post APEASE<br><br>Green text – consensus about inclusion<br>Brown text – some amendments made<br>Red text – rejected and not taken further | Framing within the Socio-ecological model |
|---|---|---|---|---|---|---|---|---|
| | | | | | | | | 55. Community and individual change |
| | (Potential) PrEP users find it difficult to discuss PrEP with others because they believe that this is private information. | (Potential) PrEP users find it easy discussing PrEP with others because they have an open approach to their sexual health. | **Example 1**<br>*"If it [PrEP] ever comes up in conversation, I might be okay saying that I am taking it, but other than that, I don't feel like it's something that I need to disclose with everyone."* (PrEP user)<br><br>**Example 2**<br>*"If I was meeting someone new, I definitely would bring it [PrEP] in to the conversation. 'Cause either way when I think about it, it's…they need to know that I have told them that I am doing this, that they know that* | Intentions<br><br>Beliefs about consequences | Persuasion<br><br>Education<br><br>Modelling | 13.2 Framing /reframing<br><br>6.3 Information about others' approval<br><br>6.1 Demonstration of the behaviour<br><br>16.3 Vicarious consequences | 56. SHCPs and CBO staff could persuade potential PrEP users and PrEP users to talk openly about PrEP and their PrEP status by emphasising that sex is between two or more people and that sexual partners will approve of being informed about what measures are in place to protect against HIV (13.2, 6.3).<br><br>57. Employ various methods (e.g. a PrEP storyline in a popular TV show, well-known and diverse HIV/PrEP activists) to demonstrate to potential PrEP users and PrEP users how to talk openly about PrEP and their PrEP status (6.1) and showcase positive outcomes (e.g. portrays honesty, helps to build a | 56. Organisational change, community change, individual change<br><br>57. Community change |

**Supplementary files: Matrices of the intervention development analyses used to theorise key barriers and facilitators to develop evidence-based and theoretically informed recommendations for future interventions to improve PrEP awareness and access**

| Priority areas for intervention | Barriers | Facilitators | Indicative quotes | TDF domains | Intervention functions | Potential BCTs | Agreed final recommendations for those considering implementing PrEP at scale- post APEASE<br><br>Green text – consensus about inclusion<br>Brown text – some amendments made<br>Red text – rejected and not taken further | Framing within the Socio-ecological model |
|---|---|---|---|---|---|---|---|---|
| | | | *I am honest. You know, whichever way they take that. It's up to them really."* (PrEP user) | | | | foundation for a trusting relationship) (16.3). | |
| | (Potential) PrEP users find it difficult to discuss PrEP with others because they are concerned about, or have experienced, PrEP-related stigma. | (Potential) PrEP users find it easy to discuss PrEP with others because they believe that PrEP is a responsible and positive means of reducing the likelihood of acquiring HIV and want to educate others. | *"I haven't dared tell her [my sister] I'm on PrEP, she'd go through the roof. Because she'd turn round and say, why should I be paying for your sex life?"* (PrEP user) | Beliefs about consequences<br><br>Environmental context and resources | Persuasion<br><br>Education<br><br>Enablement<br><br>Training | 13.2 Framing /reframing<br><br>5.1 Information about health consequences<br><br>5..3 Information about social and environmental consequences<br><br>5.6 Information about emotional consequences<br><br>9.1 Credible source<br><br>3.1 Social support (unspecified) | 58. Ensure PrEP information and communications (e.g. national patient information booklet, SHCP-and CBO staff-client interactions, posters in SHS waiting areas and consultation rooms and CBO settings, SHS, CBO, and HIV/PrEP activists' websites and social media, marketing campaign) address PrEP-related stigma, for example, by adopting 'needs-based' terminology rather than focusing on 'risk', presenting PrEP as a responsible choice and positive means of reducing the likelihood of acquiring HIV (13.2), and detailing the economic and wider benefits and value of PrEP for the healthcare system, communities, and individuals (5.1, 5.3, 5.6, 9.1) | **58.Community change** |

**Supplementary files: Matrices of the intervention development analyses used to theorise key barriers and facilitators to develop evidence-based and theoretically informed recommendations for future interventions to improve PrEP awareness and access**

| Priority areas for intervention | Barriers | Facilitators | Indicative quotes | TDF domains | Intervention functions | Potential BCTs | Agreed final recommendations for those considering implementing PrEP at scale- post APEASE<br><br>Green text – consensus about inclusion<br>Brown text – some amendments made<br>Red text – rejected and not taken further | Framing within the Socio-ecological model |
|---|---|---|---|---|---|---|---|---|
| | | | | | | 4.1 Instruction on how to perform the behaviour | 59. SHCPs and CBO staff should encourage and support PrEP users to have holistic conversations with important others about the meaning of PrEP (3.1), for instance, by sharing example phrases that clients could incorporate into discussions (4.1).<br><br>60. SHCPs and CBO staff should persuade PrEP users to talk about PrEP with important others by informing them of the important health, social, and emotional benefits of doing so (e.g. increase awareness and uptake of PrEP, reduce PrEP-related stigma) (5.1, 5.3, 5.6, 9.1). | 59. Organisational change, community change, individual change<br><br>60. Organisational change, community change, individual change |
| | | (Potential) PrEP users find it easy to discus PrEP with others because of the normalisation of PrEP in the | **Example 1**<br>*"I think in the gay scene it's such an open conversation and it's quite an open dialogue, on various platforms like* | Environmental context and resources | Environmental restructuring<br><br>Enablement<br><br>Education | 12.2 Restructure the social environment | 40. Public health agencies and those who provide HIV treatments should consider working with relevant CBOs to build on the activism and peer influence seen among GBMSM (12.2) and | **40. Community and interpersonal change** |

**Supplementary files: Matrices of the intervention development analyses used to theorise key barriers and facilitators to develop evidence-based and theoretically informed recommendations for future interventions to improve PrEP awareness and access**

| Priority areas for intervention | Barriers | Facilitators | Indicative quotes | TDF domains | Intervention functions | Potential BCTs | Agreed final recommendations for those considering implementing PrEP at scale- post APEASE<br><br>Green text – consensus about inclusion<br>Brown text – some amendments made<br>Red text – rejected and not taken further | Framing within the Socio-ecological model |
|---|---|---|---|---|---|---|---|---|
| | | GBMSM community (e.g. 'negative on PrEP' on dating apps and hook-up sites). | *social media conversations, the news and stuff like that as well, that I think it's definitely a lot more kind of prevalent and open for conversation on the gay scene."* (PrEP user)<br><br>**Example 2**<br>*"There's a lot more understanding…it's much more of a positive thing, where people now freely ask, are you on PrEP, are you not on PrEP. And it's a very sociably acceptable question, where, in 2017, it probably wasn't."* (SHCP) | Social influences | | 5.1 Information about health consequences<br><br>5.3 Information about social and environmental consequences<br><br>5.6 Information about emotional consequences<br><br>9.1 Credible source | consider peer-led PrEP awareness-raising and normalising interventions (5.1, 5.3, 5.6, 9.1) for other communities where there may be considerable benefits from wider uptake of PrEP.<br><br>61. Encourage shared learning among CBOs to build on the activism and peer influence seen among GBMSM, for example, mentorship for those working with people from trans, Black African, and injecting drug communities (12.2). | **61. Organisational change** |

**Supplementary files: Matrices of the intervention development analyses used to theorise key barriers and facilitators to develop evidence-based and theoretically informed recommendations for future interventions to improve PrEP awareness and access**

### Priority area 4: CBO staff raising PrEP with key communities

| Priority areas for intervention | Barriers | Facilitators | Indicative quotes | TDF domains | Intervention functions | Potential BCTs | Agreed final recommendations for those considering implementing PrEP at scale- post APEASE<br><br>Green text – consensus about inclusion<br>Brown text – some amendments made<br>Red text – rejected and not taken further | Framing within the Socio-ecological model |
|---|---|---|---|---|---|---|---|---|
| CBO staff engage key communities with PrEP | Some CBO staff find it difficult to engage key communities with PrEP because their organisations feel disenfranchised from the wider HIV sector. | | *"I almost felt there was a little bit of people trying to control the information around PrEP, and feeling that, you know, we weren't out there shouting from the rooftops about PrEP, and we should have been. So I think, for me, it was very frustrating, that sort of, three months after it."* (CBO staff working with GBMSM) | Environmental context and resources | Environmental restructuring<br><br>Enablement | 12.2 Restructure the social environment | 1. Prior to and throughout PrEP implementation, national leaders should provide a range of diverse opportunities (e.g. consultation in decision-making processes, workshops and information sharing events) for the full range of HIV stakeholders (i.e. CBOs, community members, SHCPs) to work together in partnership and in synergy bringing the unique strengths of a broad range of organisations together (12.2). These dynamics must reflect the full breadth of communities affected by HIV (e.g., black african communities, drug users) | **1. Public Policy change** |
| | CBO staff find it difficult to engage key communities with PrEP because many clients have competing needs | | *"When you look at the hierarchy of needs… people's priority is not health, it's education, employment, housing, merit, so for us to* | Environmental context and resources | Environmental restructuring<br><br>Education | 12.2 Restructure the social environment | 2. Those providing health and social care should ensure that partnerships and reciprocal referral mechanisms exist across a broad range of organisations that meet the diverse and sometimes | **2. Public Policy and organisational change** |

**Supplementary files: Matrices of the intervention development analyses used to theorise key barriers and facilitators to develop evidence-based and theoretically informed recommendations for future interventions to improve PrEP awareness and access**

| Priority areas for intervention | Barriers | Facilitators | Indicative quotes | TDF domains | Intervention functions | Potential BCTs | Agreed final recommendations for those considering implementing PrEP at scale- post APEASE<br><br>Green text – consensus about inclusion<br>Brown text – some amendments made<br>Red text – rejected and not taken further | Framing within the Socio-ecological model |
|---|---|---|---|---|---|---|---|---|
| | that rival HIV prevention (e.g. addiction, poor mental health, poverty, refugee/ asylum issues). | | *continually think health is actually a priority for people, you're going to get it wrong."* (CBO staff working with Black African communities) | Beliefs about consequenc es | Persuasion | 3.1 Social support (unspecified)<br><br>5.1 Beliefs about health consequences<br><br>5.3 Information about social and environmental consequences<br><br>5.6 Information about emotional consequences<br><br>9.1 Credible source | competing needs of those who may benefit from PrEP (12.2).<br><br>3. CBOs should establish good connections with other specialist services (e.g. addictions, mental health, refugee and asylum seeker support) (12.2) that CBO staff could signpost and/or directly refer clients to, for appropriate expert support for their other needs (3.1).<br><br>4. CBO staff persuade clients to prioritise HIV prevention among competing needs via a range of educational methods (e.g. posters, national patient information booklets, interactional workshops at diverse community venues, drop-in information sessions, peer-led support groups) that inform about the health, social, and emotional effects of HIV and benefits of PrEP and emphasise | **3. Public Policy and organisational change**<br><br>**4. Community change** |

**Supplementary files: Matrices of the intervention development analyses used to theorise key barriers and facilitators to develop evidence-based and theoretically informed recommendations for future interventions to improve PrEP awareness and access**

| Priority areas for intervention | Barriers | Facilitators | Indicative quotes | TDF domains | Intervention functions | Potential BCTs | Agreed final recommendations for those considering implementing PrEP at scale- post APEASE<br><br>Green text – consensus about inclusion<br>Brown text – some amendments made<br>Red text – rejected and not taken further | Framing within the Socio-ecological model |
|---|---|---|---|---|---|---|---|---|
| | | | | | | | the importance of maintaining good sexual health for overall health and wellbeing and addressing other life priorities (5.1, 5.3, 5.6, 9.1). | |
| | Some CBO staff find it difficult to engage key communities with PrEP because funding cuts and structures stifle innovation and curtail cross-sector partnerships (e.g. service level agreements, competition for dwindling resources). | CBO staff find it easy to engage key communities with PrEP because they have established effective partnerships with other CBOs, clinical teams, and commissioners of CBO services. | *"There's ongoing dialogue between ourselves and our frontline workers, and the people who commission the service. So, you know, we will flag up things that maybe we feel we could be doing more of, and when it comes to revisiting the SLA, assuming that we've been listened to, understood and agreed with, that stuff might well find its way into the SLA."* (CBO staff working with GBMSM) | Environmental context and resources | Environmental restructuring<br><br>Enablement | 12.2 Restructure the social environment<br><br>3.2 Social support (practical) | 5. Government and public health agencies need to be aware that PrEP implementation demands a coordinated network and connections between all stakeholders (e.g. SHS, CBOs, community representatives) and drive effective partnership work (12.2), for example, ring-fence funds for community engagement and clinical work (3.2)<br><br>6. Prior to and throughout PrEP implementation, commissioners of CBO services should ensure their funding mechanisms do not stifle innovation and cross-sector partnership, but encourage it (12.2). | **5. Public Policy change**<br><br>**6. Public Policy change** |

**Supplementary files: Matrices of the intervention development analyses used to theorise key barriers and facilitators to develop evidence-based and theoretically informed recommendations for future interventions to improve PrEP awareness and access**

| Priority areas for intervention | Barriers | Facilitators | Indicative quotes | TDF domains | Intervention functions | Potential BCTs | Agreed final recommendations for those considering implementing PrEP at scale- post APEASE<br><br>Green text – consensus about inclusion<br>Brown text – some amendments made<br>Red text – rejected and not taken further | Framing within the Socio-ecological model |
|---|---|---|---|---|---|---|---|---|
| | | | | | | | 7. Establish and actively maintain open lines of communication between CBO staff and commissioners of CBO services to ensure shared understandings of priorities (12.2). | **7. Organisational and interpersonal change** |
| | Some CBO staff find it difficult to engage key communities with PrEP because they struggle to adapt their previous skills to PrEP. | Some CBO staff find it easy to engage key communities with PrEP because they have longstanding adaptable expertise in HIV prevention. | **Example 1**<br>*"The arrival of PrEP in the mix has created a bit of a cultural shift in terms of our health promotion messages, messaging to MSM. It's added something very new and very significant into the mix of an offering that, for a very long time, was all about condom use, was all about barrier protection. And, you know, I think it's fair to say that the NHS and the third sector are going through a period of kind of cultural change in* | Environmental context and resources<br><br>Skills<br><br>Professional role and identity | Environmental restructuring<br><br>Training<br><br>Persuasion | 12.2 Restructure the social environment<br><br>6.1 Demonstration of the behaviour<br><br>6.2 Social comparison<br><br>13.2 Framing /reframing<br><br>15.1 Verbal persuasion about capability | 8. Prior to and throughout PrEP implementation, national leaders should acknowledge that engaging a wide range of communities with PrEP will bring diverse challenges. Working with MSM, for example, may well be easier than working with some Black African communities or people who inject drugs. Ensure mechanisms are in place to foster shared learning across diverse organisations serving a range of communities (12.2). Provide opportunities for CBO staff, across and within diverse organisations, to develop critical HIV literacy skills and discuss approaches to HIV risk reduction in the PrEP era | **8. Public Policy change** |

**Supplementary files: Matrices of the intervention development analyses used to theorise key barriers and facilitators to develop evidence-based and theoretically informed recommendations for future interventions to improve PrEP awareness and access**

| Priority areas for intervention | Barriers | Facilitators | Indicative quotes | TDF domains | Intervention functions | Potential BCTs | Agreed final recommendations for those considering implementing PrEP at scale- post APEASE<br><br>Green text – consensus about inclusion<br>Brown text – some amendments made<br>Red text – rejected and not taken further | Framing within the Socio-ecological model |
|---|---|---|---|---|---|---|---|---|
| | | | *order to be able to accommodate this new element of messaging, whilst not abandoning all the safer sex stuff that we've all been living with and promoting for a very, very long time. I mean, the landscape has changed enormously."* (CBO staff working with GBMSM)<br><br>**Example 2**<br>*"Prevention has always been part of what we do, really, in essence. You know, we go to some, we do some workshops, talking about prevention, and how to use condoms, and all that. So PrEP is just another tool, besides condoms, and the other things that are used to* | | | | and share successful methods for engaging clients with PrEP via peer learning and reflection (6.1, 6.2).<br><br>9. Within PrEP training, for CBO staff expressing particular concerns, introduce PrEP as an extension to their longstanding HIV prevention work (i.e. rather than as a standalone health promotion intervention) (13.2) and reassure them that their previous experience is still relevant and valuable for successfully engaging clients in conversations about PrEP (15.1). | 9. interpersonal change |

**Supplementary files: Matrices of the intervention development analyses used to theorise key barriers and facilitators to develop evidence-based and theoretically informed recommendations for future interventions to improve PrEP awareness and access**

| Priority areas for intervention | Barriers | Facilitators | Indicative quotes | TDF domains | Intervention functions | Potential BCTs | Agreed final recommendations for those considering implementing PrEP at scale- post APEASE<br><br>Green text – consensus about inclusion<br>Brown text – some amendments made<br>Red text – rejected and not taken further | Framing within the Socio-ecological model |
|---|---|---|---|---|---|---|---|---|
| | | | *prevent HIV."* (CBO staff working with GBMSM) | | | | | |
| | CBO staff find it difficult to engage key communities with PrEP because some groups of clients have very low levels of sexual health and HIV literacy and struggle to talk about their sexuality, sexual health, and HIV prevention needs (e.g. because of cultural stigmas attached to sex and a history of being underserved in sex education). | CBO staff find it easy to engage key communities with PrEP because they have high levels of cultural competency in delivering sexual health and HIV prevention interventions. | *"I guess it's a more informal relationship that we would have with those communities and we're perceived very often as more approachable, particularly within the MSM work. Because very often, it is gay or bisexual men who are delivering that service, so there's a point of identification there with the service user. They'll very often open up to a third sector health promotion worker in a way that they won't to NHS staff."* (CBO staff working with GBMSM) | Environmental context and resources<br><br>Knowledge<br><br>Skills | Environmental restructuring<br><br>Education<br><br>Training | 12.2 Restructure the social environment<br><br>5.1 Information about health consequences<br><br>4.1 Instruction on how to perform the behaviour<br><br>6.1 Demonstration of the behaviour<br><br>8.1 Behavioural practice/rehearsal<br><br>2.2 Feedback on behaviour(s) | 10. Governments should make age-appropriate and comprehensive relationships and sex education taught as part of the curriculum for children and young people at all levels of schooling, with fact-oriented and non-judgemental content that addresses the sexual health, social, and cultural needs of LGBTQ+ and Black African communities (12.2), incorporating issues such as PrEP as HIV prevention (5.1).<br><br>11. Where possible, CBOs should recruit staff and volunteers from among the communities they serve (12.2). CBO staff should articulate and share their cultural competency (e.g. use acceptable terminology) and ensure peer | **10. Public Policy change**<br><br>11.Community, Organisational and interpersonal change |

**Supplementary files: Matrices of the intervention development analyses used to theorise key barriers and facilitators to develop evidence-based and theoretically informed recommendations for future interventions to improve PrEP awareness and access**

| Priority areas for intervention | Barriers | Facilitators | Indicative quotes | TDF domains | Intervention functions | Potential BCTs | Agreed final recommendations for those considering implementing PrEP at scale- post APEASE<br><br>Green text – consensus about inclusion<br>Brown text – some amendments made<br>Red text – rejected and not taken further | Framing within the Socio-ecological model |
|---|---|---|---|---|---|---|---|---|
| | | | | | | | learning and training of new staff in this regard (4.1, 6.1, 8.1, 2.2).<br><br>12. CBO staff could deliver programmatic work (e.g. webinars, interactional workshops at diverse community venues, social media over a considered period of time to establish trust) and engage in outreach work (e.g. at trans facing events) (12.2) to enhance and broaden engagement within the communities they serve to increase sexual health and HIV literacy (5.1) and normalise and encourage talking about sexuality, sexual health, and HIV prevention in everyday contexts (6.1), especially among people from trans and Black African communities. | **12. Community change** |

**Supplementary files: Matrices of the intervention development analyses used to theorise key barriers and facilitators to develop evidence-based and theoretically informed recommendations for future interventions to improve PrEP awareness and access**

**Supplementary files: Matrices of the intervention development analyses used to theorise key barriers and facilitators to develop evidence-based and theoretically informed recommendations for future interventions to improve PrEP awareness and access**

### Priority area 5: SHCPs acquiring knowledge of PrEP and PreP processes

| Priority areas for intervention | Barriers | Facilitators | Indicative quotes | TDF domains | Intervention functions | Potential BCTs | Agreed final recommendations for those considering implementing PrEP at scale- post APEASE<br><br>Green text – consensus about inclusion<br>Brown text – some amendments made<br>Red text – rejected and not taken further | Framing within the socioecological model |
|---|---|---|---|---|---|---|---|---|
| SHCPs acquire knowledge of PrEP and PrEP processes. | SHCPs find it difficult to acquire knowledge of PrEP and PrEP processes because the quick roll-out of the PrEP programme meant that the national training materials came out after PrEP had started. | SHCPs find it easy to acquire knowledge of PrEP and PrEP processes because they receive 'official' nationally-developed PrEP training prior to PrEP roll-out. | **Example 1** *"This huge workload had suddenly been imposed on us with no extra resources and obviously people didn't really know anything about it and there was a lot of education to be done for staff but we had to do it all ourselves. The training slides that came out, came out well after the date of introduction."* (SHCP)<br><br>**Example 2** *"It's letting the clinics know even three months before a programme's rolled out to say – this is the* | Environmental context and resources<br><br>Knowledge | Environmental restructuring<br><br>Training<br><br>Education | 12.2 Restructure the social environment<br><br>5.1 Information about health consequences<br><br>5.3 Information about social and environmental consequences<br><br>5.6 Information about emotional consequences<br><br>9.1 Credible source | 13. Governments and public health agencies responsible for PrEP should ensure a well-paced timescale for PrEP implementation that allows for critical planning activities, such as working in partnership across the whole HIV sector to develop and deliver 'official' national PrEP training package (9.1), including education on the positive health, social, and emotional impacts of PrEP (5.1, 5.3, 5.6) and examples of how to deliver PrEP | **13. Public Policy change** |

**Supplementary files: Matrices of the intervention development analyses used to theorise key barriers and facilitators to develop evidence-based and theoretically informed recommendations for future interventions to improve PrEP awareness and access**

| **Priority areas for intervention** | **Barriers** | **Facilitators** | **Indicative quotes** | **TDF domains** | **Intervention functions** | **Potential BCTs** | **Agreed final recommendations for those considering implementing PrEP at scale- post APEASE**<br><br>**Green text – consensus about inclusion**<br>**Brown text – some amendments made**<br>**Red text – rejected and not taken further** | **Framing within the socioecological model** |
|---|---|---|---|---|---|---|---|---|
| | | | *training package; this is how you access it…that would have been really helpful."* (SHCP) | | | 4.1 Instruction on how to perform the behaviour<br><br>6.1 Demonstration of the behaviour<br><br>8.1 Behavioural practice/rehearsal<br><br>2.2 Feedback on behaviour | (4.1, 6.1), to prepare the workforce (12.2). Such training should also focus on enhancing the cultural competencies of all staff to work with diverse communities (4.1, 6.1, 8.1, 2.2). | |
| | SHCPs find it difficult to acquire knowledge of PrEP and PrEP processes because of limited opportunities to take up training (e.g. no slack in the system to free | SHCPs find it easy to acquire knowledge of PrEP and PrEP processes because of formal and informal training and learning opportunities at local- and national-level. | **Example 1** *"We did this within existing capacity which was already stretched. So, one of the issues with that was the actual capacity to train people and deliver PrEP and get people up to speed had to be found within the service, and* | Environmental context and resources<br><br>Knowledge | Enablement<br><br>Environmental restructuring<br><br>Education | 3.2 Social support (practical)<br><br>12.2 Restructuring the social environment | 14. Those that fund SHS should provide the resource required to match the costs of the programme (i.e. increase the budget according to predicted PrEP demand to ensure adequate staff capacity for effective | **14. Public Policy change** |

**Supplementary files: Matrices of the intervention development analyses used to theorise key barriers and facilitators to develop evidence-based and theoretically informed recommendations for future interventions to improve PrEP awareness and access**

| Priority areas for intervention | Barriers | Facilitators | Indicative quotes | TDF domains | Intervention functions | Potential BCTs | Agreed final recommendations for those considering implementing PrEP at scale- post APEASE<br><br>Green text – consensus about inclusion<br>Brown text – some amendments made<br>Red text – rejected and not taken further | Framing within the socioecological model |
|---|---|---|---|---|---|---|---|---|
| | up staff, few clients on PrEP). | | *actually even the simple fact of releasing people from clinics to do any training was a challenge."* (SHCP)<br><br>**Example 2***"There was a West of Scotland Managed Clinical Network masterclass. It was open to, you know, health boards to participate, so the three doctors from here went along to that. And that was just giving, obviously, background, what the criteria would be, what the background is, what the evidence has shown regarding PrEP."* (SHCP) | | | 5.1 Information about health consequences<br><br>4.1 Instructions on how to perform the behaviour<br><br>6.1 Demonstration of the behaviour<br><br>5.3 Information about social and environmental consequences | implementation and scale-up) in the initial months of national rollout (3.2). A business case produced that also outlines the health benefits of PrEP (5.1) and potential future savings of PrEP implementation within the healthcare system (i.e. more cost-effective than spending on HIV treatment) (5.3) could be helpful in this regard (9.1).<br><br>15. Offer a range of formal and informal opportunities for SHCPs to train and learn about PrEP, for example, through working closely with CBOs and at local- | <br><br><br><br><br><br><br><br><br><br><br><br><br><br><br><br>**15. Interpersonal change** |

**Supplementary files: Matrices of the intervention development analyses used to theorise key barriers and facilitators to develop evidence-based and theoretically informed recommendations for future interventions to improve PrEP awareness and access**

| Priority areas for intervention | Barriers | Facilitators | Indicative quotes | TDF domains | Intervention functions | Potential BCTs | Agreed final recommendations for those considering implementing PrEP at scale- post APEASE<br><br>Green text – consensus about inclusion<br>Brown text – some amendments made<br>Red text – rejected and not taken further | Framing within the socioecological model |
|---|---|---|---|---|---|---|---|---|
| | | | | | | | (e.g. journal clubs, team meetings, study days, shadowing), regional- (e.g. clinical network arrangements), and national-level (e.g. shared learning events) (12.2).<br><br>16. National coordinated interdisciplinary PrEP training should include inter-disciplinary online PrEP learning resources for SHCPs which can be broken down into short modules on specific topics and spread out over a period of time (5.1, 4.1). These could be aligned with CPD for many job roles. | <br><br><br><br><br><br><br><br>**16. Interpersonal change** |

**Supplementary files: Matrices of the intervention development analyses used to theorise key barriers and facilitators to develop evidence-based and theoretically informed recommendations for future interventions to improve PrEP awareness and access**

| Priority areas for intervention | Barriers | Facilitators | Indicative quotes | TDF domains | Intervention functions | Potential BCTs | Agreed final recommendations for those considering implementing PrEP at scale- post APEASE<br><br>Green text – consensus about inclusion<br>Brown text – some amendments made<br>Red text – rejected and not taken further | Framing within the socioecological model |
|---|---|---|---|---|---|---|---|---|
| | | | | | | | 17. Introduce a shadowing scheme across different SHSs to enable SHCPs from SHS with few PrEP users to become familiar with PrEP processes (12.2, 6.1). | 17. Organisational and interpersonal change |
| | SHCPs find it difficult to acquire knowledge of PrEP and PrEP processes because of | SHCPs find it easy to acquire knowledge of PrEP and PrEP processes because supporting | **Example 1***"It's like, right, okay, we've changed the protocol to six monthly. And they go like, hang on a moment, what? So everyone was doing* | Environmental context and resources | Environmental restructuring<br><br>Enablement<br><br>Education | 12.2 Restructure the social environment<br><br>7.1 Prompts/cues | 18. Ensure that a range of formal (e.g. team meetings, study days) and informal (e.g. huddles, email) opportunities are | **18. Organisational change** |

**Supplementary files: Matrices of the intervention development analyses used to theorise key barriers and facilitators to develop evidence-based and theoretically informed recommendations for future interventions to improve PrEP awareness and access**

| Priority areas for intervention | Barriers | Facilitators | Indicative quotes | TDF domains | Intervention functions | Potential BCTs | Agreed final recommendations for those considering implementing PrEP at scale- post APEASE<br><br>Green text – consensus about inclusion<br>Brown text – some amendments made<br>Red text – rejected and not taken further | Framing within the socioecological model |
|---|---|---|---|---|---|---|---|---|
| | changes to how PrEP is delivered. | documents are available. | *three-monthly scripts last week. Yes, so we've had another meeting, we've had a discussion, so it's changed. So…it's just a case of because we're working at warp speed people do suddenly feel they get a bit of whiplash every now and then."* (SHCP)<br><br>**Example 2***"It helped having, you know, useful documents we could go to especially in the early days when it all seemed so new. So, having good sort of supporting documentation knowing if somebody came into a drop-in clinic what the process was."* (SHCP) | Memory, attention, and decision processes<br><br>Knowledge | | | available to cascade changes to PrEP processes to all relevant SHCPs (12.2).<br><br>19. Create and update paper-based or electronic checklists/ proformas, crib sheets, and flowcharts (e.g. based on a formal protocol) that SHCPs can use to remind themselves of PrEP processes (7.1) | **19. Organisational change** |

**Supplementary files: Matrices of the intervention development analyses used to theorise key barriers and facilitators to develop evidence-based and theoretically informed recommendations for future interventions to improve PrEP awareness and access**

**Supplementary files: Matrices of the intervention development analyses used to theorise key barriers and facilitators to develop evidence-based and theoretically informed recommendations for future interventions to improve PrEP awareness and access**

### Priority area 6: SHCPs engage potential PrEP users with PrEP

| Priority areas for intervention | Barriers | Facilitators | Indicative quotes | TDF domains | Intervention functions | Potential BCTs | Agreed final recommendations for those considering implementing PrEP at scale- post APEASE<br><br>Green text – consensus about inclusion<br>Brown text – some amendments made<br>Red text – rejected and not taken further | Framing within the socioecological model |
|---|---|---|---|---|---|---|---|---|
| SHCPs engage potential PrEP users with PrEP | | SHCPs find it easy to engage potential PrEP users with PrEP because nationally-developed patient information booklets (e.g. i-Base PrEP in Scotland, Know about PrEP tool) and other supporting documents (e.g. quick guides) help them to introduce PrEP and structure | **Example 1**<br>*"I'd probably pick up one of the PrEP leaflets and go through it, because it's quite a good leaflet."* (SHCP)<br><br>**Example 2**<br>*"They already had some literature waiting, so we just opened one of them, went through a couple of things…"* (PrEP user) | Environmental context and resources<br><br>Memory, attention, and decision processes | Enablement<br><br>Training | 7.1 Prompt/cues<br><br>4.1 Instructions on how to perform the behaviour | 20. Drawing on experiences within other national settings where PrEP has already been implemented, co-develop a range of resources that address a range of PrEP literacy needs (e.g. fact sheet, PrEP provider pocket guide, national patient information booklets, 'how to' scripts) to help SHCPs introduce PrEP and structure PrEP conversations (7.1, 4.1). Such resources should ideally be co-produced by a range of diverse organisations and the | 20. Community change |

**Supplementary files: Matrices of the intervention development analyses used to theorise key barriers and facilitators to develop evidence-based and theoretically informed recommendations for future interventions to improve PrEP awareness and access**

| Priority areas for intervention | Barriers | Facilitators | Indicative quotes | TDF domains | Intervention functions | Potential BCTs | Agreed final recommendations for those considering implementing PrEP at scale- post APEASE<br><br>Green text – consensus about inclusion<br>Brown text – some amendments made<br>Red text – rejected and not taken further | Framing within the socioecological model |
|---|---|---|---|---|---|---|---|---|
| | | PrEP conversations | | | | | communities who will use them | |
| | SHCPs find it difficult to engage potential PrEP users with PrEP because many vulnerable MSM and people from other HIV-affected communities (e.g. Black Africans, trans people, people who inject drugs) do not appear to be attending SHS | SHCPs find it easy to engage potential PrEP users with PrEP because clients present to SHS already knowledgeable about or self-seeking PrEP (i.e. they have high levels of HIV and PrEP literacy) | **Example 1**<br>*"We've seen no transgender people…it's been exclusively MSM. So yeah, I guess, is the information getting out to, particularly, transgender groups locally, that they would eligible for it as well. So, perhaps that's something that we need to look at locally, because they're definitely there, but they're not coming to our service."* (SHCP)<br><br>**Example 2**<br>*"I think a lot of men already know about PrEP now so if they're not coming in asking for it* | Environmental context and resources | Environmental restructuring<br><br>Enablement<br><br>Education | 12.2 Restructure the social environment<br><br>3.1 Social support (unspecified)<br><br>5.1 Information about health consequences<br><br>5.3 Information about social and environmental consequences<br><br>5.6 Information about emotional consequences | 21. Government, public health agencies, and those commissioning and providing PrEP services should foster partnerships across SHS and CBOs (12.2) and ensure awareness and locations of PrEP services are widely disseminated (3.1)<br><br>22. Partnership work between SHS, CBOs, and community representatives should co-produce tailored resources to raise awareness of HIV and PrEP and provide PrEP information (5.1, 5.3, 5.6) in the languages, tones, and formats most | 21. Policy change<br><br>**22. Community change** |

**Supplementary files: Matrices of the intervention development analyses used to theorise key barriers and facilitators to develop evidence-based and theoretically informed recommendations for future interventions to improve PrEP awareness and access**

| Priority areas for intervention | Barriers | Facilitators | Indicative quotes | TDF domains | Intervention functions | Potential BCTs | Agreed final recommendations for those considering implementing PrEP at scale- post APEASE<br><br>Green text – consensus about inclusion<br>Brown text – some amendments made<br>Red text – rejected and not taken further | Framing within the socioecological model |
|---|---|---|---|---|---|---|---|---|
| | | | *when you mention it they go, yes, I've heard all about that or my friend's on it or when I'm on Grindr people say, on PrEP. So, it's way more out there and it's not quite such a huge conversation."* (SHCP) | | | 13.2 Framing /reframing<br><br>2.7 Feedback on outcome(s) of behaviour<br><br>12.1 Restructure the physical environment | accessible by and acceptable to the intended audience, including advice (e.g. clear statements and nuanced examples) regarding the eligibility criteria (13.2, 2.7). Ensure these resources are disseminated and distributed through culturally appropriate means (12.1)<br><br>23. Work with SHCPs within each SHS to work with CBO staff and HIV/PrEP activists to engage with wider communities who are *not* attending SHS (e.g. Black Africans, trans people, people who inject drugs) (12.2) such people should | **23 Individual and organisational change** |

**Supplementary files: Matrices of the intervention development analyses used to theorise key barriers and facilitators to develop evidence-based and theoretically informed recommendations for future interventions to improve PrEP awareness and access**

| Priority areas for intervention | Barriers | Facilitators | Indicative quotes | TDF domains | Intervention functions | Potential BCTs | Agreed final recommendations for those considering implementing PrEP at scale- post APEASE<br><br>Green text – consensus about inclusion<br>Brown text – some amendments made<br>Red text – rejected and not taken further | Framing within the socioecological model |
|---|---|---|---|---|---|---|---|---|
| | | | | | | | review and share uptake data and co-ordinate referral pathways into SHS and support colleague's training | |
| | SHCPs find it difficult to engage potential PrEP users with PrEP because PrEP adds considerable extra time to already typically lengthy and complex consultations in time-pressed clinics, compounded by the coinciding introduction of the HPV vaccination | SHCPs find it easy to engage potential PrEP users with PrEP because there is a shared understanding among colleagues that PrEP takes extra time (e.g. no pressure to complete the pre-PrEP workup in the initial consultation, not viewed as skiving if taking a long time with a client) | **Example 1**<br>*"PrEP is another thing to try and add into an already kind of lengthening consultation. So…we used to see people pretty quickly because you were doing A, B, and C, now we're having to spend longer with patients because we're doing A, B, C, D, PrEP and HPV."* (SHCP)<br><br>**Example 2**<br>*"We didn't want our nursing staff to feel they were being criticised if, you know, I didn't do this* | Environmental context and resources<br><br>Social influences | Enablement<br><br>Environmental restructuring | 3.2 Social support (practical)<br><br>12.1 Restructure the physical environment<br><br>12.2 Restructure the social environment | 14. Those that fund SHS should provide the resource required to match the costs of the programme (i.e. increase the budget according to predicted PrEP demand to ensure adequate staff capacity for effective implementation and scale-up) in the initial months of national rollout (3.2). A business case that also outlines the health benefits of PrEP (5.1) and potential future savings of PrEP implementation within the healthcare system (i.e. more cost- | **14. Public Policy change** |

**Supplementary files: Matrices of the intervention development analyses used to theorise key barriers and facilitators to develop evidence-based and theoretically informed recommendations for future interventions to improve PrEP awareness and access**

| Priority areas for intervention | Barriers | Facilitators | Indicative quotes | TDF domains | Intervention functions | Potential BCTs | Agreed final recommendations for those considering implementing PrEP at scale- post APEASE<br><br>Green text – consensus about inclusion<br>Brown text – some amendments made<br>Red text – rejected and not taken further | Framing within the socioecological model |
|---|---|---|---|---|---|---|---|---|
| | programme for MSM | | *or, you know...At least if they got the discussion going that was the important thing. If the patient came back for the follow-up appointment, then that'd been a success."* (SHCP) | | | | effective than spending on HIV treatment) (5.3) could be helpful in this regard (9.1)<br><br>24. Government and public health agencies should ensure that the roll-out of PrEP does not coincide with the introduction of other programmes (12.1, 12.2) or if this is unavoidable / it is preferable to make a major change through introducing two innovations at once (i.e. so one period of disruption not two), that appropriate resources are devoted to measured service reorganisation (3.2) | 24. Policy change |

**Supplementary files: Matrices of the intervention development analyses used to theorise key barriers and facilitators to develop evidence-based and theoretically informed recommendations for future interventions to improve PrEP awareness and access**

| Priority areas for intervention | Barriers | Facilitators | Indicative quotes | TDF domains | Intervention functions | Potential BCTs | Agreed final recommendations for those considering implementing PrEP at scale- post APEASE<br><br>Green text – consensus about inclusion<br>Brown text – some amendments made<br>Red text – rejected and not taken further | Framing within the socioecological model |
|---|---|---|---|---|---|---|---|---|
| | | | | | | | 25. Those involved in the organisation of services should facilitate discussions (e.g. in team meetings, huddles) on what is realistically achievable within consultations, acknowledge that PrEP does take extra time and agree minimum expectations (e.g. having an initial discussion) to ensure shared understandings among SHCPs and a supportive working environment (12.2)<br><br>26. SHS should explore and provide innovative ways of scheduling appointments with built-in flexibility to respond to | 25 . Policy change<br><br>26 . Policy change |

**Supplementary files: Matrices of the intervention development analyses used to theorise key barriers and facilitators to develop evidence-based and theoretically informed recommendations for future interventions to improve PrEP awareness and access**

| Priority areas for intervention | Barriers | Facilitators | Indicative quotes | TDF domains | Intervention functions | Potential BCTs | Agreed final recommendations for those considering implementing PrEP at scale- post APEASE<br><br>Green text – consensus about inclusion<br>Brown text – some amendments made<br>Red text – rejected and not taken further | Framing within the socioecological model |
|---|---|---|---|---|---|---|---|---|
| | | | | | | | long standing health inequalities in health and HIV literacy and varying client need (e.g. longer discussions about PrEP and wider sexual health issues) (12.2) | |
| | SHCPs find it difficult to engage potential PrEP users with PrEP because they fear that they might stigmatise or offend people from communities other than MSM by highlighting their risks for HIV | SHCPs find it easy to engage potential PrEP users with PrEP because they PrEP is a natural extension to conversations already taking place in MSM consultations | **Example 1**<br>*"I think it is probably work that needs to come from the community, from them. Rather than for us to say, you do release that you're at a higher risk because all your partners were Black African men, you know. That, in itself, can put people on the back foot. But if it's somebody in the community, that says, look actually, traditionally, we have* | Beliefs about consequences<br><br>Skills<br><br>Professional role and identity | Education<br><br>Training<br><br>Persuasion | 5.3 Information about social and environmental consequences<br><br>6.1 Demonstration of the behaviour<br><br>4.1 Instruction on how to perform the behaviour | 27. SHS should work closely with CBOs to educate and train all SHCPs to sensitively highlight risks for HIV and motivate them to consider PrEP (for example, among trans and Black African clients (5.3, 6.1), taking account of their specific sexual health cultures and acceptable terminology)<br><br>28. Close SHS-CBO partnership work could deliver a list of culturally | 27. Community change<br><br>28. Community change |

**Supplementary files: Matrices of the intervention development analyses used to theorise key barriers and facilitators to develop evidence-based and theoretically informed recommendations for future interventions to improve PrEP awareness and access**

| Priority areas for intervention | Barriers | Facilitators | Indicative quotes | TDF domains | Intervention functions | Potential BCTs | Agreed final recommendations for those considering implementing PrEP at scale- post APEASE<br><br>Green text – consensus about inclusion<br>Brown text – some amendments made<br>Red text – rejected and not taken further | Framing within the socioecological model |
|---|---|---|---|---|---|---|---|---|
| | | | *equal rates of HIV…"* (SHCP)<br><br>**Example 2**<br>*"The skills were there, anyway, because we were working with MSM. We've always done health promotion, anyway. So, I just felt it [PrEP] was an extension of a role we already had, we were always doing health promotion, we were always doing screening, we were always talking about incubation periods, safer sex, condom use."* (SHCP) | | | 6.2 Social comparison<br><br>2.2 Feedback on behaviour<br><br>2.3 Self-monitoring of behaviour<br><br>13.2 Framing /reframing<br><br>15.1 Verbal persuasion about capabilities | appropriate phrases to help SHCPs flexibly tailor their language when highlighting HIV risks among non-MSM clients (e.g. people who inject drugs, trans people, people from Black African communities) (4.1)<br><br>29. Provide opportunities for SHCPs to share successful conversational approaches for highlighting HIV risks to trans and Black African clients via peer learning and peer reflection (6.1, 6.2) and reflect on their skills during clinical supervision and annual appraisals (2.2, 2.3) | 29. Organisational and interpersonal change |

**Supplementary files: Matrices of the intervention development analyses used to theorise key barriers and facilitators to develop evidence-based and theoretically informed recommendations for future interventions to improve PrEP awareness and access**

| Priority areas for intervention | Barriers | Facilitators | Indicative quotes | TDF domains | Intervention functions | Potential BCTs | Agreed final recommendations for those considering implementing PrEP at scale- post APEASE<br><br>Green text – consensus about inclusion<br>Brown text – some amendments made<br>Red text – rejected and not taken further | Framing within the socioecological model |
|---|---|---|---|---|---|---|---|---|
| | | | | | | | 30. Within PrEP training for SHCPs, introduce PrEP as a natural extension to conversations that are already taking place in MSM consultations (i.e. rather than as a standalone health promotion intervention) (13.2) and reassure them that their previous experience is still relevant and valuable for successfully engaging clients in conversations about PrEP (15.1) | 30. Organisational and interpersonal change |

**Supplementary files: Matrices of the intervention development analyses used to theorise key barriers and facilitators to develop evidence-based and theoretically informed recommendations for future interventions to improve PrEP awareness and access**

### Priority area 7: SCHPs provide access to PrEP

| Priority areas for intervention | Barriers | Facilitators | Indicative quotes | TDF domains | Intervention functions | Potential BCTs | Agreed final recommendations for those considering implementing PrEP at scale- post APEASE<br><br>Green text – consensus about inclusion<br>Brown text – some amendments made<br>Red text – rejected and not taken further | Framing within the socioecological model |
|---|---|---|---|---|---|---|---|---|
| SHS provide access to PrEP | SHS find it difficult to provide access to PrEP because of resource issues arising from the lack of dedicated PrEP capacity, pre-existing clinic pressures, very high demand, and the cumulative effect of PrEP | SHS would find it easy to provide access to PrEP because PrEP is available in other settings | **Example 1**<br>*"Obviously you have to have the resource, and we still don't have any additional resource. It's staffing time… there's additional tests being done, and the consultations take longer. So, yeah, the manpower, but it's also the administrative time as well, involved in that, when test results come through, they need to be managed, so* | Environmental context and resources | Enablement<br><br>Environmental restructuring | 3.2 Social support (practical)<br><br>12.1 Restructure the physical environment<br><br>12.2 Restructure the social environment | 14. Those that fund SHS should provide the resource required to match the innovation (i.e. increase the budget according to predicted PrEP demand to ensure adequate staff capacity for effective implementation and scale-up) in the initial months of national rollout (3.2). A business case produced by senior HIV clinicians that also outlines the health benefits of PrEP (5.1) and potential future savings of PrEP implementation within the healthcare system (i.e. more cost-effective than | **14. Public policy change** |

**Supplementary files: Matrices of the intervention development analyses used to theorise key barriers and facilitators to develop evidence-based and theoretically informed recommendations for future interventions to improve PrEP awareness and access**

| Priority areas for intervention | Barriers | Facilitators | Indicative quotes | TDF domains | Intervention functions | Potential BCTs | Agreed final recommendations for those considering implementing PrEP at scale- post APEASE<br><br>Green text – consensus about inclusion<br>Brown text – some amendments made<br>Red text – rejected and not taken further | Framing within the socioecological model |
|---|---|---|---|---|---|---|---|---|
| | | | *it's all of those things, definitely not just the drugs."* (SHCP)<br><br>**Example 2**<br>*"Rather than somebody always having to come in every three months, they could do the tests at home. And, as long as we have negative results, we can continue prescriptions. So, I guess that might be something that might be helpful in the future, as more and more patients go on PrEP. I guess also, for your more straightforward* | | | | spending on HIV treatment) (5.3) could be helpful in this regard (9.1)<br><br>62. Consider alternative service models to make PrEP available via a range of settings, including all SHS (e.g. local hubs and satellites, as well as central services), remote care (e.g. ePrEP clinic, phone consultations), community venues (e.g. outreach clinics), and non-sexual health-specific health services (e.g. reproductive health clinics, GP surgery), with agreed pathways for non-complex PrEP users and those with additional medical complexity (12.1, 12.2) | <br><br><br>62. Organisational change |

**Supplementary files: Matrices of the intervention development analyses used to theorise key barriers and facilitators to develop evidence-based and theoretically informed recommendations for future interventions to improve PrEP awareness and access**

| **Priority areas for intervention** | **Barriers** | **Facilitators** | **Indicative quotes** | **TDF domains** | **Intervention functions** | **Potential BCTs** | **Agreed final recommendations for those considering implementing PrEP at scale- post APEASE**<br><br>**Green text – consensus about inclusion**<br>**Brown text – some amendments made**<br>**Red text – rejected and not taken further** | **Framing within the socioecological model** |
|---|---|---|---|---|---|---|---|---|
| | | | *patients, is the ability for other services such as, I guess primary care, to do some of the follow up, and for us to maybe see them every year."* (SHCP) | | | | | |
| | SHS find it difficult to provide access to PrEP because the timescale for PrEP implementation was rushed and did not allow much scope for planning | SHS find it easy to provide access to PrEP because of a collective commitment to improving sexual health meaning that increasing access to PrEP was prioritised (e.g. available via urgent care, absorbed into drop-ins, MSM clinics now PrEP clinics) | **Example 1**<br>*"It was a tiny bit chaotic, because the timescale was not of my choosing and seemed to be, and I need to ascend this bit, seemed to be to be overtly political, because there was a real drive to make Scotland first. And that's fine, I'm very committed to the idea. But, it meant we had a really* | Environmental context and resources<br><br>Beliefs about consequences | Environmental restructuring<br><br>Training<br><br>Enablement<br><br>Education<br><br>Persuasion | 12.1 Restructure the physical environment<br><br>12.2 Restructure the social environment<br><br>9.1 Credible source<br><br>4.1 Instruction on how to perform the behaviour | 63. Governments and public health agencies responsible for PrEP should ensure a well-paced timescale for PrEP implementation that allows for critical planning activities, such as estimating the likely demand for PrEP, conducting a full service review to determine capacity and how PrEP will fit into existing practices, and working in partnership across the whole HIV sector | **63. Public policy change** |

**Supplementary files: Matrices of the intervention development analyses used to theorise key barriers and facilitators to develop evidence-based and theoretically informed recommendations for future interventions to improve PrEP awareness and access**

| Priority areas for intervention | Barriers | Facilitators | Indicative quotes | TDF domains | Intervention functions | Potential BCTs | Agreed final recommendations for those considering implementing PrEP at scale- post APEASE<br><br>Green text – consensus about inclusion<br>Brown text – some amendments made<br>Red text – rejected and not taken further | Framing within the socioecological model |
|---|---|---|---|---|---|---|---|---|
| | | | *difficult task to try and pull together some Scotland relevant training materials for something that hadn't been delivered anywhere else, in a really short timescale and offer support to small boards particularly, but also in larger boards, who had no familiarity with these medicines."* (SHCP)<br><br>**Example 2**<br>*"Around about 7 to 8 per cent of all of our urgent care activity, which should be largely for* | | | 6.1 Demonstration of the behaviour<br><br>8.1 Behavioural practice/rehearsal<br><br>2..2 Feedback on behaviour<br><br>5.1 Information about health consequences<br><br>5.3 Information about social and environmental consequences<br><br>5.6 Information about emotional consequences<br><br>9.1 Credible source | to develop and deliver an 'official' national PrEP training package (9.1), including examples of how to deliver PrEP services (4.1, 6.1), to prepare the workforce (12.1, 12.2). Such training should also focus on enhancing the cultural competencies of all staff to work with diverse communities (4.1, 6.1, 8.1, 2.2)<br><br>64. Ensure that those responsible for organising SHS prioritise access to PrEP by educating them about the economic and wider benefits and value of PrEP for the healthcare system, local SHS, communities, and individual clients (e.g. arrange talks from leading | **64. Public Policy change** |

**Supplementary files: Matrices of the intervention development analyses used to theorise key barriers and facilitators to develop evidence-based and theoretically informed recommendations for future interventions to improve PrEP awareness and access**

| Priority areas for intervention | Barriers | Facilitators | Indicative quotes | TDF domains | Intervention functions | Potential BCTs | Agreed final recommendations for those considering implementing PrEP at scale- post APEASE<br><br>Green text – consensus about inclusion<br>Brown text – some amendments made<br>Red text – rejected and not taken further | Framing within the socioecological model |
|---|---|---|---|---|---|---|---|---|
| | | | *people with new symptoms, is now given over to assessment and prescribing of PrEP…that's how we've set it up, to try and increase access to PrEP, 'cause we think of it as a good thing."* (SHCP) | | | | HIV experts who are in favour of PrEP and inform of its positive health, cost/ financial, social, and emotional impacts) (5.1, 5.3, 5.6, 9.1) | |
| | SHS find it difficult to provide access to PrEP because they are unable to release staff for PrEP-related training due to resource issues | SHS find it easy to provide access to PrEP because of development and investment in the role of nurses, enabling them to work to agreed protocols and undertake non-medical prescribing | *Example 1*<br>*"They want to train, [illegible] we want to train th[illegible] but there's just not slack in the service at moment, until we get more staff." (SHC[illegible]*<br><br>*Example 2*<br>*"What you want to d[illegible] get as many nurses prescribers as possibl[illegible]* | Environmental context and resources<br><br>Knowledge<br><br>Skills | Enablement<br><br>Education<br><br>Training | 3.2 Social support (practical)<br><br>12.2 Restructure the social environment<br><br>5.1 Information about health consequences | 14. Those that fund SHS should provide the resource required to match the cost of the programme (i.e. increase the budget according to predicted PrEP demand to ensure adequate staff capacity for effective implementation and scale-up) in the initial months of national rollout (3.2). A business case that | **14. Public Policy and organisational change** |

**Supplementary files: Matrices of the intervention development analyses used to theorise key barriers and facilitators to develop evidence-based and theoretically informed recommendations for future interventions to improve PrEP awareness and access**

| Priority areas for intervention | Barriers | Facilitators | Indicative quotes | TDF domains | Intervention functions | Potential BCTs | Agreed final recommendations for those considering implementing PrEP at scale- post APEASE<br><br>Green text – consensus about inclusion<br>Brown text – some amendments made<br>Red text – rejected and not taken further | Framing within the socioecological model |
|---|---|---|---|---|---|---|---|---|
| | | | *make PrEP more effic and more cost effec in a service. 'Cause nu led services will hands down. So if service was planning and had time, I would get all your nurses to through a… some kin non-medical prescrib Or have a PGD ready t they will be confiden use." (SHCP)* | | | 4.1 Instruction on how to perform the behaviour | outlines the health benefits of PrEP (5.1) and potential future savings of PrEP implementation within the healthcare system (i.e. more cost-effective than spending on HIV treatment) (5.3) could be helpful in this regard (9.1)<br><br>65. Facilitate and sustain a respectful team-oriented culture that values multidisciplinary working and that develops and uses the knowledge and skills of all team members to the best effect (12.2)<br><br>66. Invest in the development of the role of nurses (3.2), for example, agree a timescale over which all nurses within the | 65.Organisational and interpersonal change<br><br>.<br><br>**66. Organisational change** |

**Supplementary files: Matrices of the intervention development analyses used to theorise key barriers and facilitators to develop evidence-based and theoretically informed recommendations for future interventions to improve PrEP awareness and access**

| Priority areas for intervention | Barriers | Facilitators | Indicative quotes | TDF domains | Intervention functions | Potential BCTs | Agreed final recommendations for those considering implementing PrEP at scale- post APEASE<br><br>Green text – consensus about inclusion<br>Brown text – some amendments made<br>Red text – rejected and not taken further | Framing within the socioecological model |
|---|---|---|---|---|---|---|---|---|
| | | | | | | | SHS will complete a course in non-medical prescribing (12.2) and/or, in the interim period, develop written instructions, within a legislative framework, that allow nurses to supply PrEP without a prescription or an instruction from a prescriber (e.g. patient group direction) (4.1) | |
| | SHS find it difficult to provide access to PrEP because PrEP adds considerable extra time to already typically lengthy and complex consultations in | SHS find it easy to provide access to PrEP because PrEP consultations become more streamlined over time (e.g. as SHCPs feel more comfortable with the process, more clients | **Example 1**<br>*"What we found was that the patients who were attending, particularly if they were new, for PrEP were taking up so much more time than their allocated half hour slot, and* | Environmental context and resources<br><br>Behavioural regulation<br><br>Knowledge | Environmental restructuring<br><br>Enablement<br><br>Training<br><br>Education | 12.1 Restructure the physical environment<br><br>12.2 Restructure the social environment<br><br>3.2 Social support (practical) | 24. Government and public health agencies should ensure that the roll-out of PrEP does not coincide with the introduction of other programmes (12.1, 12.2) or if this is unavoidable / it is preferable to make a major change through introducing two | 24. Policy change |

**Supplementary files: Matrices of the intervention development analyses used to theorise key barriers and facilitators to develop evidence-based and theoretically informed recommendations for future interventions to improve PrEP awareness and access**

| Priority areas for intervention | Barriers | Facilitators | Indicative quotes | TDF domains | Intervention functions | Potential BCTs | Agreed final recommendations for those considering implementing PrEP at scale- post APEASE<br><br>Green text – consensus about inclusion<br>Brown text – some amendments made<br>Red text – rejected and not taken further | Framing within the socioecological model |
|---|---|---|---|---|---|---|---|---|
| | time-pressed clinics, compounded by the coinciding introduction of the HPV vaccination programme for MSM | present as PrEP literate) | *that was then having a negative impact on the other patients who were booked into that clinic."* (SHCP)<br><br>**Example 2**<br>*"When we first started, I was having longer appointments and actually, after a few months, once I'd got to grips with that, I could go back and say, well actually I think we could see maybe a few more people in the PrEP clinic and just because we're better now, we've got our spiel, we've* | | | 6.1 Demonstration of the behaviour<br><br>4.1 Instruction on how to perform the behaviour<br><br>5.1 Information about health consequences | innovations at once (i.e. so one period of disruption not two), that appropriate resources are devoted to measured service reorganisation (3.2).<br><br>67. During initial roll-out, pilot a staggered approach to introducing a full service to enable staff to learn about engaging with patients effectively, shadowing each other and honing efficient consultations.<br><br>operationalise PrEP via specific clinics (12.1) to enable a core team of SHCPs to quickly build their skills and familiarity with PrEP processes and then | **67. Organisational change** |

**Supplementary files: Matrices of the intervention development analyses used to theorise key barriers and facilitators to develop evidence-based and theoretically informed recommendations for future interventions to improve PrEP awareness and access**

| Priority areas for intervention | Barriers | Facilitators | Indicative quotes | TDF domains | Intervention functions | Potential BCTs | Agreed final recommendations for those considering implementing PrEP at scale- post APEASE<br><br>Green text – consensus about inclusion<br>Brown text – some amendments made<br>Red text – rejected and not taken further | Framing within the socioecological model |
|---|---|---|---|---|---|---|---|---|
| | | | *got our way of doing it."* (SHCP) | | | | introduce a shadowing scheme where SHCPs that are new to PrEP have the opportunity to observe more experienced SHCPs 'in action' before delivering PrEP themselves (12.2, 6.1).<br><br>68. Consider developing scripts as a foundation for SHCPs to succinctly and accurately discuss PrEP with clients (4.1).<br><br>69. Develop and implement a range of awareness raising strategies to enhance PrEP literacy among groups at increased need of PrEP (e.g. provide information via SHS, CBO, | <br><br><br><br><br><br><br><br>**68. Organisational change**<br><br><br><br>**69. Community change** |

**Supplementary files: Matrices of the intervention development analyses used to theorise key barriers and facilitators to develop evidence-based and theoretically informed recommendations for future interventions to improve PrEP awareness and access**

| Priority areas for intervention | Barriers | Facilitators | Indicative quotes | TDF domains | Intervention functions | Potential BCTs | Agreed final recommendations for those considering implementing PrEP at scale- post APEASE<br><br>Green text – consensus about inclusion<br>Brown text – some amendments made<br>Red text – rejected and not taken further | Framing within the socioecological model |
|---|---|---|---|---|---|---|---|---|
| | | | | | | | and HIV/PrEP activists' websites, community champions and social media, posters in CBO, SHS, and other health settings, CBO outreach work, message blasts on dating apps and hook-up sites, marketing campaigns) (5.1). | |
| | | SHS find it easy to provide access to PrEP because good IT systems and shared learning from a strong nationally coordinated PrEP programme facilitate service innovation and adaptation to issues of time, increasing demand, and different user needs (e.g. emphasis | *"The protocols are always being changed, as more information comes through. We do want the nurses to be able to do the majority of the straightforward, non-complicated patients, and everything is set up for that…which will free up the doctors,* | Environmental context and resources<br><br>Social influences | Environmental restructuring<br><br>Enablement<br><br>Modelling | 12.2 Restructure the social environment<br><br>3.1 Social support (unspecified)<br><br>3.2 Social support (practical)<br><br>6.1 Demonstration of the behaviour | 70. Where possible, implement PrEP via a nationally coordinated programme and use local, regional, and national infrastructures for peer support to facilitate opportunities for iterative and shared learning on optimal PrEP service delivery models (e.g. email, 'phone a friend', discussion forums, workshops, PrEP Leads | **70. Public Policy change** |

**Supplementary files: Matrices of the intervention development analyses used to theorise key barriers and facilitators to develop evidence-based and theoretically informed recommendations for future interventions to improve PrEP awareness and access**

| Priority areas for intervention | Barriers | Facilitators | Indicative quotes | TDF domains | Intervention functions | Potential BCTs | Agreed final recommendations for those considering implementing PrEP at scale- post APEASE<br><br>Green text – consensus about inclusion<br>Brown text – some amendments made<br>Red text – rejected and not taken further | Framing within the socioecological model |
|---|---|---|---|---|---|---|---|---|
| | | on triage, prescribing PrEP before the pre-assessment test results are back, matching staff skills to client complexity) | *to see the more complex patients. Other things have changed. So, to begin with, the patients who were prescribed PrEP were given a month to start with. Now we can give them three months, which makes it an awful lot easier.”* (SHCP) | | | 6.2 Social comparison<br><br>12.1 Restructure the physical environment<br><br>2.7 Feedback on outcome(s) of behaviour | meetings, clinical network arrangements) (12.2, 3.1, 3.2, 6.1, 6.2).<br><br>71. Devise a system to monitor the PrEP programme (e.g. collect data on PrEP uptake and any associated waiting time, characteristics of PrEP seekers/users, STI and HIV rates) (12.1) and review data on a regular basis to inform service planning (2.7).<br><br>72. Introduce an effective triage system to ensure optimal flow of clients through the SHS (i.e. they are directed to the most appropriate SHCP) and efficient use of scarce resources (12.1, 12.2). | **71. Organisational change**<br><br>**72. Organisational change** |

**Supplementary files: Matrices of the intervention development analyses used to theorise key barriers and facilitators to develop evidence-based and theoretically informed recommendations for future interventions to improve PrEP awareness and access**

| Priority areas for intervention | Barriers | Facilitators | Indicative quotes | TDF domains | Intervention functions | Potential BCTs | Agreed final recommendations for those considering implementing PrEP at scale- post APEASE<br><br>Green text – consensus about inclusion<br>Brown text – some amendments made<br>Red text – rejected and not taken further | Framing within the socioecological model |
|---|---|---|---|---|---|---|---|---|
| | | | | | | | 73. In line with WHO guidelines, PrEP providers should move to routine use of point of care rapid HIV tests and starting clients on PrEP on the same day that they present to SHS, with the exception of special circumstances (e.g. exposure to HIV in the last 72 hours, signs/symptoms of acute HIV infection, known renal issues) and so long as they agree to be contacted and return to see a SHCP if any of the baseline test results require action, confirmation, or treatment (12.1).<br><br>74. SHS should consider having a nurse-led care pathway for non-complex | 29 Policy and organisatonal change<br><br>74. Organisational change |

**Supplementary files: Matrices of the intervention development analyses used to theorise key barriers and facilitators to develop evidence-based and theoretically informed recommendations for future interventions to improve PrEP awareness and access**

| Priority areas for intervention | Barriers | Facilitators | Indicative quotes | TDF domains | Intervention functions | Potential BCTs | Agreed final recommendations for those considering implementing PrEP at scale- post APEASE<br><br>Green text – consensus about inclusion<br>Brown text – some amendments made<br>Red text – rejected and not taken further | Framing within the socioecological model |
|---|---|---|---|---|---|---|---|---|
| | | | | | | | PrEP users and a doctor-led care pathway for those with additional medical complexity (12.2). | |

**Supplementary files: Matrices of the intervention development analyses used to theorise key barriers and facilitators to develop evidence-based and theoretically informed recommendations for future interventions to improve PrEP awareness and access**

### Priority area 8: Potential PrEP user accesses sexual health services and PrEP care

| Priority areas for intervention | Barriers | Facilitators | Indicative quotes | TDF domains | Intervention functions | Potential BCTs | Agreed final recommendations for those considering implementing PrEP at scale- post APEASE<br><br>Green text – consensus about inclusion<br>Brown text – some amendments made<br>Red text – rejected and not taken further | Framing within socioeconomic model |
|---|---|---|---|---|---|---|---|---|
| Potential PrEP users access PrEP | | Potential PrEP users find it easy to access PrEP because they already attend SHS so receive advance notice or are proactively contacted by a SHCP (i.e. because they appear to meet the eligibility criteria) about PrEP availability | *"When we started to talk about PrEP, you know, we would have shared that information with those patients and then just kept them up-to-date with things and, you know, eventually then once it was available these were patients then that were prescribed."* (SHCP) | Environmental context and resources<br><br>Knowledge | Enablement<br><br>Education | 3.1 Social support (unspecified)<br><br>5.1 Information about health consequences<br><br>2.7 Feedback on outcome(s) of behaviour<br><br>12.2 Restructure the social environment | 75. SHCPs should keep clients informed about PrEP availability (e.g. coming soon, provide the date for roll-out) at the SHS during consultations (3.1).<br><br>51. At initial PrEP roll-out and routinely (e.g. quarterly) thereafter, SHCPs could run a report on the IT system to identify clients who (likely) meet the eligibility criteria but have not had a PrEP discussion and attempt to make contact via email, SMS, or phone to inform them about the health benefits of PrEP (5.1), its | **75. Interpersonal change**<br><br>**51. Organisational change** |

**Supplementary files: Matrices of the intervention development analyses used to theorise key barriers and facilitators to develop evidence-based and theoretically informed recommendations for future interventions to improve PrEP awareness and access**

| Priority areas for intervention | Barriers | Facilitators | Indicative quotes | TDF domains | Intervention functions | Potential BCTs | Agreed final recommendations for those considering implementing PrEP at scale- post APEASE<br><br>Green text – consensus about inclusion<br>Brown text – some amendments made<br>Red text – rejected and not taken further | Framing within socioeconomic model |
|---|---|---|---|---|---|---|---|---|
| | | | | | | | availability at the SHS (3.1), and their potential eligibility (2.7) and offer a rapid appointment (12.2). | |
| | Potential PrEP users find it difficult to access PrEP because there are limited options for where (e.g. at some not all SHS, located far away), when (e.g. inconvenient time slots), and how (e.g. by appointment, set up to be delivered in male only or MSM clinics) they can access specialist | Potential PrEP users find it easy to access PrEP because there is flexibility in where (e.g. at all SHS, in other more valued / acceptable settings), when (e.g. extended opening hours), and how (e.g. via drop-in clinics, by appointment) they can access PrEP | **Example 1**<br>*"Where PrEP is offered is limiting people of colour, of the African continent, to go. Because most of them don't want to go to a sexual health clinic, most of them, when they have problems, they go to their GP. And…most of the young women I know, that are in my circle, they do go to the reproductive health clinic. And that [PrEP] is not being offered in the reproductive health clinic. They are offered condoms, why are we not offering them PrEP."* (CBO | Environmental context and resources | Environmental restructuring<br><br>Enablement | 12.1 Restructure the physical environment<br><br>12.2 Restructure the social environment<br><br>3.1 Social support (unspecified) | 62. Consider alternative service models to make PrEP available to potential PrEP users via a range of settings, including all SHS (e.g. local hubs and satellites, as well as central services), remote care (e.g. ePrEP clinic, phone consultations), community venues (e.g. outreach clinics), and non-sexual health-specific health services (e.g. reproductive health clinics, GP surgery), with agreed pathways for non-complex PrEP users and those with additional medical complexity (12.1, 12.2). | **62. Organisational change** |

**Supplementary files: Matrices of the intervention development analyses used to theorise key barriers and facilitators to develop evidence-based and theoretically informed recommendations for future interventions to improve PrEP awareness and access**

| Priority areas for intervention | Barriers | Facilitators | Indicative quotes | TDF domains | Intervention functions | Potential BCTs | Agreed final recommendations for those considering implementing PrEP at scale- post APEASE<br><br>Green text – consensus about inclusion<br>Brown text – some amendments made<br>Red text – rejected and not taken further | Framing within socioeconomic model |
|---|---|---|---|---|---|---|---|---|
| | PrEP start appointments | | *staff working with Black African communities)*<br><br>**Example 2**<br>*"In an ideal world people could just, they could come to any clinic and they would be seen and they wouldn't have to go to a specialist, we would call it a specialist clinic at the moment, and they would be able to come in, we have a lot of drop-in clinics and they would just be able to have everything done, get their PrEP." (SHCP)* | | | | 76. Establish PrEP as routine clinical practice within SHS and implement through regular drop-in clinics, in addition to booked appointments (12.1) offering protected spaces for women for example.<br><br>77. Maximise all drop-in visits by ensuring there is sufficient waiting space, toilets, and consultation rooms (12.1) and operationalising drop-in clinics via a multidisciplinary team of SHCPs who can task-share and accommodate complex cases (12.2). | **76. Organisational change**<br><br>**77. Organisational change**<br><br>**78. Organisational change** |

**Supplementary files: Matrices of the intervention development analyses used to theorise key barriers and facilitators to develop evidence-based and theoretically informed recommendations for future interventions to improve PrEP awareness and access**

| Priority areas for intervention | Barriers | Facilitators | Indicative quotes | TDF domains | Intervention functions | Potential BCTs | Agreed final recommendations for those considering implementing PrEP at scale- post APEASE<br><br>Green text – consensus about inclusion<br>Brown text – some amendments made<br>Red text – rejected and not taken further | Framing within socioeconomic model |
|---|---|---|---|---|---|---|---|---|
| | | | | | | | 78. Provide access to drop-in clinics and pre-bookable appointments on mid-week evenings and at weekends to suit contemporary lifestyles and meet local population needs (12.1).<br><br>79. Support potential PrEP users in becoming aware of when and how they can access drop-in clinics and book and reschedule appointments for PrEP initiation (e.g. SHCPs and CBO staff provide information verbally, hand out location-specific leaflets or wallet-sized inserts, signpost to websites) (3.1). | **.79. Organisational, individual change** |
| | Potential PrEP users find it difficult to access | Potential PrEP users find it easy to access PrEP because they | **Example 1**<br>*"In a small community it's difficult, and we have* | Environmental context and resources | Environmental restructuring | 12.2 Restructure the social environment | 80. Governments and public health agencies could encourage CBOs and | 80. Policy change |

**Supplementary files: Matrices of the intervention development analyses used to theorise key barriers and facilitators to develop evidence-based and theoretically informed recommendations for future interventions to improve PrEP awareness and access**

| Priority areas for intervention | Barriers | Facilitators | Indicative quotes | TDF domains | Intervention functions | Potential BCTs | Agreed final recommendations for those considering implementing PrEP at scale- post APEASE<br><br>Green text – consensus about inclusion<br>Brown text – some amendments made<br>Red text – rejected and not taken further | Framing within socioeconomic model |
|---|---|---|---|---|---|---|---|---|
| | PrEP because of confidentiality concerns and stigma associated with SHS, especially in smaller towns and rural communities | are able to use any SHS within Scotland, including those in other Health Boards | *to respect that people will sometimes choose to access their services from elsewhere, and through time as we break the stigma around topics and issues then I think people will become, I suppose, more comfortable in using our services."* (SHCP)<br><br>**Example 2**<br>*"Going somewhere where, you know, you're less likely to be recognised, what have you, creates a wee bit of anonymity there which helps, I think, for the…for sexual health treatment. And that's why I prefer to go up to [city] for it [PrEP] as opposed to [health* | Beliefs about consequences | Enablement<br><br>Persuasion<br><br>Education | 5.1 Information about health consequences<br><br>5.3 Information about social and environmental consequences<br><br>5.6 Information about emotional consequences<br><br>13.2 Framing/ reframing<br><br>12.1 Restructure the physical environment<br><br>3.2 Social support (practical) | SHS to work together to establish a set of criteria that could be used to affirm organisational attainment of cultural competencies and assure potential service users of confidentiality (e.g. similar to investors in people) (12.2).<br><br>81. Use a multi-method approach (e.g. CBO staff-client interactions, SHS, CBO, and HIV/PrEP activists' websites and social media, posters in non-sexual health services, sex and relationships education) to normalise SHS attendance by presenting sex and sexual health as integral rather than peripheral to overall | **81. Policy and community change .** |

**Supplementary files: Matrices of the intervention development analyses used to theorise key barriers and facilitators to develop evidence-based and theoretically informed recommendations for future interventions to improve PrEP awareness and access**

| Priority areas for intervention | Barriers | Facilitators | Indicative quotes | TDF domains | Intervention functions | Potential BCTs | Agreed final recommendations for those considering implementing PrEP at scale- post APEASE<br><br>Green text – consensus about inclusion<br>Brown text – some amendments made<br>Red text – rejected and not taken further | Framing within socioeconomic model |
|---|---|---|---|---|---|---|---|---|
| | | | *board] area. And that probably helps me to keep going because there's limited chance for that embarrassment happening if I was to bump in to someone."* (PrEP user) | | | | health and wellbeing (5.1, 5.3, 5.6), framing attending SHS as a responsible behaviour with favourable outcomes for individuals, their sexual partner(s), and wider communities (e.g. peace of mind, timely treatment if they receive a positive test result, prevent onward transmission of STIs/ HIV) (5.1, 5.3, 5.6), and encouraging potential PrEP users to view SHS attendance like any other routine health appointment (13.2).<br><br>82. Educate potential PrEP users, for example, via CBO staff-client interactions, SHS, CBO, and HIV/PrEP activists' websites and social media, posters in | <br><br><br><br><br><br><br><br><br><br><br><br><br><br><br><br><br>82. Community change |

**Supplementary files: Matrices of the intervention development analyses used to theorise key barriers and facilitators to develop evidence-based and theoretically informed recommendations for future interventions to improve PrEP awareness and access**

| Priority areas for intervention | Barriers | Facilitators | Indicative quotes | TDF domains | Intervention functions | Potential BCTs | Agreed final recommendations for those considering implementing PrEP at scale- post APEASE<br><br>Green text – consensus about inclusion<br>Brown text – some amendments made<br>Red text – rejected and not taken further | Framing within socioeconomic model |
|---|---|---|---|---|---|---|---|---|
| | | | | | | | non-sexual health services, and sex and relationships education, about what to expect when they attend SHS and reassure them that *all* HCPs, including SHCPs, have a duty of confidentiality and that the information they provide will only be used to ensure they receive the most appropriate care (5.1, 5.3).<br><br>83. Co-locate SHS with other healthcare services to allow clients, especially those in smaller towns and rural areas, some discretion about the reason for their attendance (12.1).<br><br>84. Enable potential PrEP users to access PrEP via SHS outside their Health Board | 83. **Organisational change**<br><br>84. Policy and organisational change |

**Supplementary files: Matrices of the intervention development analyses used to theorise key barriers and facilitators to develop evidence-based and theoretically informed recommendations for future interventions to improve PrEP awareness and access**

| Priority areas for intervention | Barriers | Facilitators | Indicative quotes | TDF domains | Intervention functions | Potential BCTs | Agreed final recommendations for those considering implementing PrEP at scale- post APEASE<br><br>Green text – consensus about inclusion<br>Brown text – some amendments made<br>Red text – rejected and not taken further | Framing within socioeconomic model |
|---|---|---|---|---|---|---|---|---|
| | | | | | | | area (i.e. permit countrywide attendance) and agree reimbursement of PrEP medication costs between Health Boards (12.1, 3.2). | |
| | Potential PrEP users find it difficult to access PrEP because they, or their important others (e.g. peers, sexual partners, friends, family), have previous negative experiences of SHS and the wider healthcare system (e.g. institutional racism, | Potential PrEP users find it easy to access PrEP because of signposting/referral and encouragement from important others (e.g. peers, sexual partners, friends, family), CBO staff, and other HCPs (e.g. GPs) | **Example 1**<br>*"If trans people hear these stories…because obviously trans people talk to each other, trans people are hugely active online, there are big groups… you know, if you follow [organisation] for any length of time, you'll get blow-by-blow detail of all the horrific things that happen to people at sexual health clinics and inappropriate questions and…it's a minefield for trans people."* (CBO staff | Environmental context and resources<br><br>Social influences | Environmental restructuring<br><br>Enablement<br><br>Education | 12.2 Restructure the social environment<br><br>12.1 Restructure the physical environment<br><br>1.2 Problem solving<br><br>5.3 Information about social and environmental consequences<br><br>5.6 Information about emotional consequences | 85. Governments and public health agencies could encourage SHS and CBOs to work together to establish a set of criteria that could be used to affirm organisational attainment of cultural competencies (e.g. similar to investors in people) (12.2). In this way, many barriers to accessing PrEP are systematically reduced.<br><br>86. Working in collaboration with CBOs, SHS should explore the previous experiences of | **85. Organisational change**<br><br>**86. Organisational change** |

**Supplementary files: Matrices of the intervention development analyses used to theorise key barriers and facilitators to develop evidence-based and theoretically informed recommendations for future interventions to improve PrEP awareness and access**

| Priority areas for intervention | Barriers | Facilitators | Indicative quotes | TDF domains | Intervention functions | Potential BCTs | Agreed final recommendations for those considering implementing PrEP at scale- post APEASE<br><br>Green text – consensus about inclusion<br>Brown text – some amendments made<br>Red text – rejected and not taken further | Framing within socioeconomic model |
|---|---|---|---|---|---|---|---|---|
| | homophobia, transphobia) | | working with trans people)<br><br>**Example 2**<br>*"I've done that with PrEP with my own friends, you know…people come and ask you about it and they say, well, tell me about it, tell me about your experiences with it and what do you do, and how do I get. I give them that information. And as I said, five have actually acted on it."* (PrEP user) | | | 5.1 Information about health consequences<br><br>9.1 Credible source<br><br>2.3 Self-monitoring of behaviour<br><br>3.1 Social support (unspecified) | SHS and the wider healthcare system among a diverse sample of clients from Black African, MSM, and trans communities to understand what was handled well and why and what could be improved upon and how (1.2). Ensure that findings are disseminated widely (e.g. through SHS, CBO, and HIV/ PrEP activist networks, websites, and social media) and state clearly any subsequent changes that will be made to improve service provision as a direct result of the work (12.1, 12.2).<br><br>87. Facilitate and actively maintain (e.g. via training, huddles, clinical | 87. **Organisational change** |

**Supplementary files: Matrices of the intervention development analyses used to theorise key barriers and facilitators to develop evidence-based and theoretically informed recommendations for future interventions to improve PrEP awareness and access**

| Priority areas for intervention | Barriers | Facilitators | Indicative quotes | TDF domains | Intervention functions | Potential BCTs | Agreed final recommendations for those considering implementing PrEP at scale- post APEASE<br><br>Green text – consensus about inclusion<br>Brown text – some amendments made<br>Red text – rejected and not taken further | Framing within socioeconomic model |
|---|---|---|---|---|---|---|---|---|
| | | | | | | | supervision, reflective practice, signage and changes to settings) a clearly warm, welcoming, and friendly atmosphere wherein SHCPs communicate with clients in a non-judgemental manner, using inclusive, sex- and PrEP-positive, and destigmatising language to establish trust and ensure an open dialogue (12.2, 5.3).<br><br>88. Establish and actively maintain a positive organisational culture (12.2) by educating SHCPs in a wholistic understanding of sexual health and wellbeing, equalities, racism, heterosexism, and trans- | **88. Organisational change** |

**Supplementary files: Matrices of the intervention development analyses used to theorise key barriers and facilitators to develop evidence-based and theoretically informed recommendations for future interventions to improve PrEP awareness and access**

| Priority areas for intervention | Barriers | Facilitators | Indicative quotes | TDF domains | Intervention functions | Potential BCTs | Agreed final recommendations for those considering implementing PrEP at scale- post APEASE<br><br>Green text – consensus about inclusion<br>Brown text – some amendments made<br>Red text – rejected and not taken further | Framing within socioeconomic model |
|---|---|---|---|---|---|---|---|---|
| | | | | | | | and homophobia (5.3, 5.6, 5.1), reflecting a wholistic approach in the SHS values and mission statement and including as a core competency for professional conduct, and providing opportunities for regular reflective practice on mindfully not stigmatising groups or individuals (2.3).<br><br>89. SHS should assure potential PrEP users that the SHS is a welcoming, safe, and non-judgemental space through co-produced (e.g. with CBO staff, community representatives) culturally appropriate literature (e.g. posters, national patient information booklets) in | <br><br><br><br><br><br><br><br><br><br><br><br>89. **Organisational change** |

**Supplementary files: Matrices of the intervention development analyses used to theorise key barriers and facilitators to develop evidence-based and theoretically informed recommendations for future interventions to improve PrEP awareness and access**

| Priority areas for intervention | Barriers | Facilitators | Indicative quotes | TDF domains | Intervention functions | Potential BCTs | Agreed final recommendations for those considering implementing PrEP at scale- post APEASE<br><br>Green text – consensus about inclusion<br>Brown text – some amendments made<br>Red text – rejected and not taken further | Framing within socioeconomic model |
|---|---|---|---|---|---|---|---|---|
| | | | | | | | SHS waiting areas and consultation rooms and other settings (e.g. at CBOs, GP) and online information (e.g. via SHS websites and social media) (12.2).<br><br>54. SHCPs and CBO staff should encourage clients to discuss PrEP with important others by informing them of the important health, social, and emotional benefits of doing so (e.g. increase awareness and uptake of PrEP, reduce PrEP-related stigma) (5.1, 5.3, 5.6, 9.1) and help to facilitate PrEP conversations by asking clients to identify potential barriers to talking about PrEP and selecting | **54.** Organisational change, community change, Individual change |

**Supplementary files: Matrices of the intervention development analyses used to theorise key barriers and facilitators to develop evidence-based and theoretically informed recommendations for future interventions to improve PrEP awareness and access**

| Priority areas for intervention | Barriers | Facilitators | Indicative quotes | TDF domains | Intervention functions | Potential BCTs | Agreed final recommendations for those considering implementing PrEP at scale- post APEASE<br><br>Green text – consensus about inclusion<br>Brown text – some amendments made<br>Red text – rejected and not taken further | Framing within socioeconomic model |
|---|---|---|---|---|---|---|---|---|
| | | | | | | | strategies to overcome these (1.2).<br><br>55. SHCPs and CBO staff could find ways of engaging and supporting PrEP champions from diverse communities to share their expertise and experiences with a wide audience of potential PrEP users (e.g. record a testimonial) (9.1).<br><br>21. Government, public health agencies, and those commissioning and providing PrEP services should foster partnerships across SHS and CBOs (12.2) and ensure awareness and locations of PrEP services are widely disseminated (3.1). | **55.interpersonal and individual change**<br><br>21. Policy change |
